# Cross-cohort mixture analysis: a data integration approach with applications on gestational age and DNA-methylation-derived gestational age acceleration metrics

**DOI:** 10.1101/2023.04.14.23288581

**Authors:** Elena Colicino, Roberto Ascari, Hachem Saddiki, Francheska Merced-Nieves, Nicolo Foppa Pedretti, Kathi Huddleston, Robert O Wright, Rosalind J Wright, Child Health Outcomes, program collaborators for Environmental influences on Child Health Outcomes*

**Affiliations:** Department of Environmental Medicine and Public Health, Icahn School of Medicine at Mount Sinai, New York, NY, USA; Department of Economics, Management and Statistics, University of Milano-Bicocca, Milan, Italy; Department of Pediatrics, Icahn School of Medicine at Mount Sinai; George Mason University, VA, USA

## Abstract

**Background:** Data integration of multiple epidemiologic studies can provide enhanced exposure contrast and statistical power to examine associations between environmental exposure mixtures and health outcomes. Extant studies have combined population studies and identified an overall mixture-outcome association, without accounting for differences across studies.

**Objective:** To extend the novel Bayesian Weighted Quantile Sum (BWQS) regression to a hierarchical framework to analyze mixtures across multiple cohorts of different sample sizes.

**Methods:** We implemented a hierarchical BWQS (HBWQS) approach that (i) aggregates sample size of multiple cohorts to calculate an overall mixture index, thereby identifying the most harmful exposure(s) across cohorts; and (ii) provides cohort-specific associations between the overall mixture index and the outcome. We showed results from six simulated scenarios including four mixture components in five and ten populations, and two real case-examples on the association between prenatal metal mixture exposure—comprising arsenic, cadmium and lead—and both gestational age and gestational age acceleration metrics.

**Results:** Results from simulated scenarios showed good empirical coverage and little bias for all parameters estimated with HBWQS. The Watanabe-Akaike information criterion (WAIC) for the HBWQS regression showed a better average performance across scenarios than the BWQS regression. HBWQS results incorporating cohorts within the national Environmental influences on Child Health Outcomes (ECHO) program from three different sites (Boston, New York City (NYC), and Virginia) showed that the environmental mixture—composed of low levels of arsenic, cadmium, and lead—was negatively associated with gestational age in NYC..

**Conclusions:** This novel statistical approach facilitates the combination of multiple cohorts and accounts for individual cohort differences in mixture analyses. Findings from this approach can be used to develop regulations, policies, and interventions regarding multiple co-occurring environmental exposures and it will maximize use of extant publicly available data.

## 1. Introduction

Environmental exposures are emerging as key factors to understanding the origin and prevention of human diseases.^1^ Although environmental exposures rarely occur in isolation, prevailing statistical approaches have traditionally analyzed them individually.^2, 3^ To accommodate the complexity of multiple co-occurring exposures and to assess their joint association with diseases, multiple mixture approaches^2, 3^ have been developed. Existing mixture approaches incorporate correlation structure among exposures, thereby limiting both a collinearity effect and standard-error inflation.^4, 5^ However, those methods fail to replicate mixture-outcome associations across cohorts,^6^ which limits their applicability to regulatory decisions.

To solve this issue, the majority of the studies have combined population studies and identified an overall mixture-outcome association, without taking into account the potential heterogeneity across key characteristics of the studies being combined.^6, 7^ A few novel statistical approaches have also proposed to identify clusters and factors commonly shared across datasets. For example, multi-study factor analyses identify factors common to all studies leveraging on joint models while the multiple dataset integration approach identifies groups of features that tend to be grouped together in multiple datasets, using a Dirichlet-multinomial allocation mixture and exploiting statistical dependencies between the datasets.^8–10^ However, those approaches identify clusters of features, but do not show the importance of individual features, thus yielding challenges in interpreting results.

Among the mixture approaches, our team developed the Bayesian Weighted Quantile Sum (BWQS) regression,^11^ which is a supervised quantile-based approach to assess the association of an outcome with multiple environmental exposures.^11–14^ All exposures are combined additively into a weighted index, with weights capturing the contribution of each exposure to the mixture.^12–14^ The BWQS regression provides simplicity of inference, easy interpretability, mixture-response association, and results that are insensitive to exposures’ outliers.^11–14^ In addition, the BWQS regression does not require *a priori* selection of the directionality of the mixture-outcome association, thus allowing flexibility of the coefficient estimates. Here we extend the framework of the BWQS regression to a hierarchical setting to accommodate mixture analyses across multiple cohorts of different sample sizes.

This work is organized in a stepwise fashion as follows: in Section 2, we reviewed the BWQS regression and introduce the hierarchical BWQS (HBWQS) regression model; in Section 3, we provide results from six simulated scenarios, also providing an accuracy measure. Section 4 deals with two real-case examples on the relationship between prenatal chemical exposures and both gestational age and gestational age acceleration metrics. Finally, Sections 5 and 6 offer a discussion and some concluding remarks, respectively.

## 2. Methods: regression model and distributions

### 2.1 The Bayesian Weighted Quantile Sum (BWQS) regression

We first review the BWQS regression as a framework for assessing the effect of a complex exposure mixture on a continuous normally distributed outcome, y_i_.^11, 15^ For each subject i=1,…,n, the BWQS regression relates the outcome to M components of the mixture **z**_i_=(z_1i_,…,z_Mi_) through a weighted index 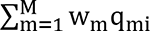, which represents the linear combination of the quantiles **q**_i_=(q_1i_,…,q_Mi_) of **z**_i_, i.e. the empirical cumulative distribution function of **z**_i_,, and with w_m_ identifying the weights mapped to the m-th mixture component (z_m_). The model can be adjusted for C relevant covariates **x**_i_=(x_1i_,…,x_Ci_) and its general form is:

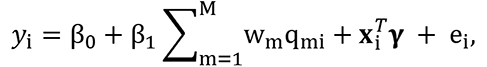

where β_0_ is the intercept, β_1_ is the regression coefficient representing the overall linear effect between the weighted index and the outcome, ****γ****= (*γ*_1_,…, *γ*_C_) represents the effects of the potential confounders on the outcome, and e_i_ is the error term distributed as a N(0,σ_y_^2^), with σ_y_^2^ being an InvGamma(0.01, 0.01). ^11, 15^ Considering a Bayesian estimation procedure, we impose a weakly informative priors for all the regression parameters **β** = (β_0_, β_1_) and ****γ**** by eliciting two independent multivariate Normal distribution with null mean vector and diagonal covariance matrix with large values (e.g. 100) on the diagonal. As the vector of weights **w** = (w_1_,…, w_M_) follows a Dirichlet prior distribution parameterized by a vector of **1**. ^11, 15^

### 2.2 The hierarchical Bayesian Weighted Quantile Sum (HBWQS) regression for multiple cohorts

We now assume J (j=1,…J) cohorts harmonized and combined in a single dataset with the goals of **1)** identifying the overall mixture index that summarizes the most harmful mixture compounds across cohorts and **2)** assessing the cohort-specific association between the overall mixture index and the outcome of interest. We define the hierarchical BWQS regression as:

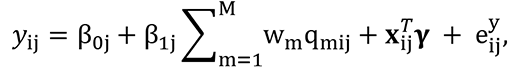

where **β_j_** = (β_0j_, β_1j_) are random terms of the j-th cohort, respectively intercept and coefficient mapped to the mixture, following independent Normal distributions centered in **µ_β_**= (µ_β0_, µ_β1_) and with variance **σ_β_**= (σ_β0_, σ_β1_), with hyperpriors **µ_β_** ∼ N(**0**, 100\***I**), σ_β0_ ∼ IG(0.01, 0.01), σ_β1_ ∼ IG(0.0.01), and **I** indicating the 2×2 diagonal matrix. In this model, the quantiles **q**_i_ are calculated using exposure levels across cohorts in order to create a common scale for the exposure mixture.

To compare the BWQS and the HBWQS regression models we can use the Watanabe-Akaike information criterion (WAIC), which is comparable to the Akaike Information Criteria for frequentist approaches. The WAIC compares models fitting the same data, and it is computed using the log-likelihood evaluated at the posterior simulations of the parameter values.^16, 17^ The WAIC closely approximates a Bayesian cross-validation due to its foundation on the series expansion of leave one out cross validation.^16, 17^

## 3. Simulation studies

In this Section, we reported the results of a simulation study involving six parametric scenarios. Namely, we simulated six scenarios including 500 synthetic datasets each. To show the stability and goodness of our models, we reported the mean estimates, the bias, the mean square error (MSE) and the empirical coverage of all parameters (**β_j_**, **w**, ****γ****, and σ_y_^2^) for all scenarios.

In the first five scenarios, we simulated each synthetic dataset comprising five cohorts, while in the sixth scenario we considered 10 cohorts. In all cohorts, we assumed to have a continuous outcome, four mixture components, which were ranked using quartiles, and three covariates. We included two continuous and one binary covariates, whose respectively coefficients were *γ* = (−0.4, 0.2, 0.5). The cohort sample sizes were assumed to be different in order to mimic the reality. In the first five scenarios, the five cohorts had the following sample sizes: 250, 200, 150, 100, and 100, while the sixth scenario included 10 cohorts of 96, 87, 96, 60, 70, 107, 56, 53, 106, and 72 observations. We assumed σ_y_^2^ =1 throughout scenarios, unless it is differently specified.

**Scenario 1.** We assumed that the mixture components contributed differently to the overall mixture **w**=[0.1,0.4,0.3,0.1] and that the random intercept and slope **β_j_** = (β_0j_, β_1j_) in the j-th cohort (j=1,..,5) were **β_1_** = (0.65, −1.7); **β_2_** = (−0.38, −0.94); **β_3_** = (0.15, −0.11); **β_4_** = (−0.98, −1.45); **β**_5_ = (1.13, −1.88).

**Scenario 2.** We assumed to have a predominant weight in the overall mixture (i.e. with higher weight w_m_ compared to the other components) and we set the mixture component weights to **w**=[0.7,0.1,0.1,0.1]. The remaining parameters were unchanged from Scenario 1.

**Scenario 3.** We evaluated the effect of having a very small weight for a specific mixture component: **w**=[0.30, 0.45, 0.24, 0.01] and random terms were unchanged from Scenario 1. **Scenario 4.** Compared to Scenario 1, we assumed that in the first cohort there was no association between the mixture and the outcome, so we had different random terms for first cohort **β_1_** = (0.65, 0), while we left unchanged the remaining random terms and weights.

**Scenario 5.** We assumed a larger model variance σ_y_ ^2^ =3.12 compared to all other scenarios, while we left unchanged all parameters from Scenario 1.

**Scenario 6**. We assumed 10 cohorts with random terms of **β_1_** = (0.65, −1.7); **β_2_** = (−0.38, −0.94); **β_3_** = (0.15, −0.11); **β_4_** = (−0.98, −1.45); **β_5_** = (1.13, −1.88), **β_6_** = (−0.55, −0.87); **β_7_** = (0.98, −0.04); **β_8_** = (0.55, −0.81); **β_9_** = (−0.18, −1.95); **β_10_** = (0.13, −0.58). We changed the variance of the model to be σ_y_^2^ =0.8. We left the weights unchanged from Scenario 1.

We implemented our models and all analyses leveraging Stan, which is a C++ library for Bayesian inference using the No-U-Turn sampler (a variant of Hamiltonian Monte Carlo), and RStan, an R interface to Stan. We illustrated the HBWQS code at https://github.com/ElenaColicino/bwqs/.

Each model included a single Markov chain of 10,000 iterations with a warm-up of 5,000 and thinning of 10. **Table S1** summarizes the truth, estimate, bias, mean square error (MSEs) and empirical coverage of the parameters across all scenarios, while **Figures S1**-**S2** showed the true value, the estimate, and the empirical coverage of all parameters.

**Table 1.** Main characteristics of study participants of the PRISM Boston (BOS), PRISM New York City (NYC) and F1000 studies.

| Variable | PRISM BOS, n = 252<br>Mean (SD)<br>or N (%) | PRISM NYC, n = 389<br>Mean (SD)<br>or N (%) | F1000, n = 708<br>Mean (SD)<br>or N (%) | p-value<sup>c</sup> |
| --- | --- | --- | --- | --- |
| **Gestational age (weeks)** | 39.17 (1.44) | 38.53 (2.45) | 39.31 (1.12) | <0.001 |
| **Gestational age acceleration metrics (weeks)** |  |  |  |  |
| RPC | 0.00 (0.88)<sup>a</sup> | -0.03 (1.00)<sup>b</sup> |  | 0.93 |
| CPC | 0.00 (0.90)<sup>a</sup> | 0.02 (1.00)<sup>b</sup> |  | 0.68 |
| refRPC | -0.07 (0.93)<sup>a</sup> | 0.07 (0.99)<sup>b</sup> |  | 0.34 |
| **Log2-transformed exposure levels (log2(µg/L))** |  |  |  |  |
| Arsenic (As) | 3.45 (1.48) | 3.42 (1.46) | 2.57 (1.58) | <0.001 |
| Cadmium (Cd) | -2.83 (1.16) | -2.20 (1.27) | -3.04 (1.38) | <0.001 |
| Lead (Pb) | -1.43 (1.24) | -0.65 (1.10) | -1.65 (0.89) | <0.001 |
| **Maternal age at birth (years)** | 31.95 (5.20) | 28.17 (6.04) | 32.48 (4.83) | <0.001 |
| **Urine creatinine (ug/mL)** | 892.14 (527.46) | 1,266.85 (762.90) | 809.45 (566.49) | <0.001 |
| **Maternal education binary** |  |  |  | <0.001 |
| Above high school | 61 (24%) | 182 (47%) | 106 (15%) |  |
| High school or less | 191 (76%) | 207 (53%) | 602 (85%) |  |
| **Maternal race/ethnicity** |  |  |  | <0.001 |
| Black/Hispanic black | 52 (21%) | 220 (57%) | 27 (3.8%) |  |
| Non-black Hispanic | 84 (33%) | 148 (38%) | 97 (14%) |  |
| Non-Hispanic non-black | 96 (38%) | 13 (3.3%) | 397 (56%) |  |
| Others | 20 (7.9%) | 8 (2.1%) | 187 (26%) |  |
| **Parity** |  |  |  | <0.001 |
| More than one | 102 (40%) | 130 (33%) | 141 (20%) |  |
| None | 54 (21%) | 127 (33%) | 225 (32%) |  |
| One | 96 (38%) | 132 (34%) | 342 (48%) |  |
| **Child sex** |  |  |  | 0.99 |
| Female | 125 (50%) | 191 (49%) | 351 (50%) |  |
| Male | 127 (50%) | 198 (51%) | 357 (50%) |  |

<sup>a</sup> n=106<sup>b</sup> n=60<sup>c</sup> p-value indicating the variable differences across populations. t-test for continuous variables and Chi-Square test for categorical variables.
RPC= robust placental clock; CPC: control placental clock; refRPC: refined RPC
SD: standard deviation

Across scenarios, all parameters showed very little bias ranging from −0.83 to 0.57 for random terms, which were in the [-1.95, 1.13] domain, and from −0.01 to 0.03 for weights, which individually were in the [0,1] domain. The larger biases for random terms were in Scenario 5 due to the large model variability (σ_y_^2^ =3.12). In general, posterior means provided reliable estimates also in terms of mean squared errors (MSEs) (**Table S1**). The MSEs were small with the exceptions of random intercepts in Scenario 5, due to the large model variability. All estimated values for weights showed an empirical coverage over 90%, while the majority (34/35 random intercepts and 32/35 random slopes) of random terms showed an empirical coverage above 90% across scenarios (**Table S1**). We also provided a summary of BWQS results in **Table S2**, showing good estimates for weights, while larger model variances compared to the actual values for five out of six simulated scenarios. To compare the results from HBWQS and BWQS on simulated data, we used the WAIC metrics, which indicate better performance with the smaller values. In our simulations, the WAIC metrics for the HBWQS regression showed a better average performance across scenarios than the BWQS regression (**Figure 1**).

**Figure 1.**
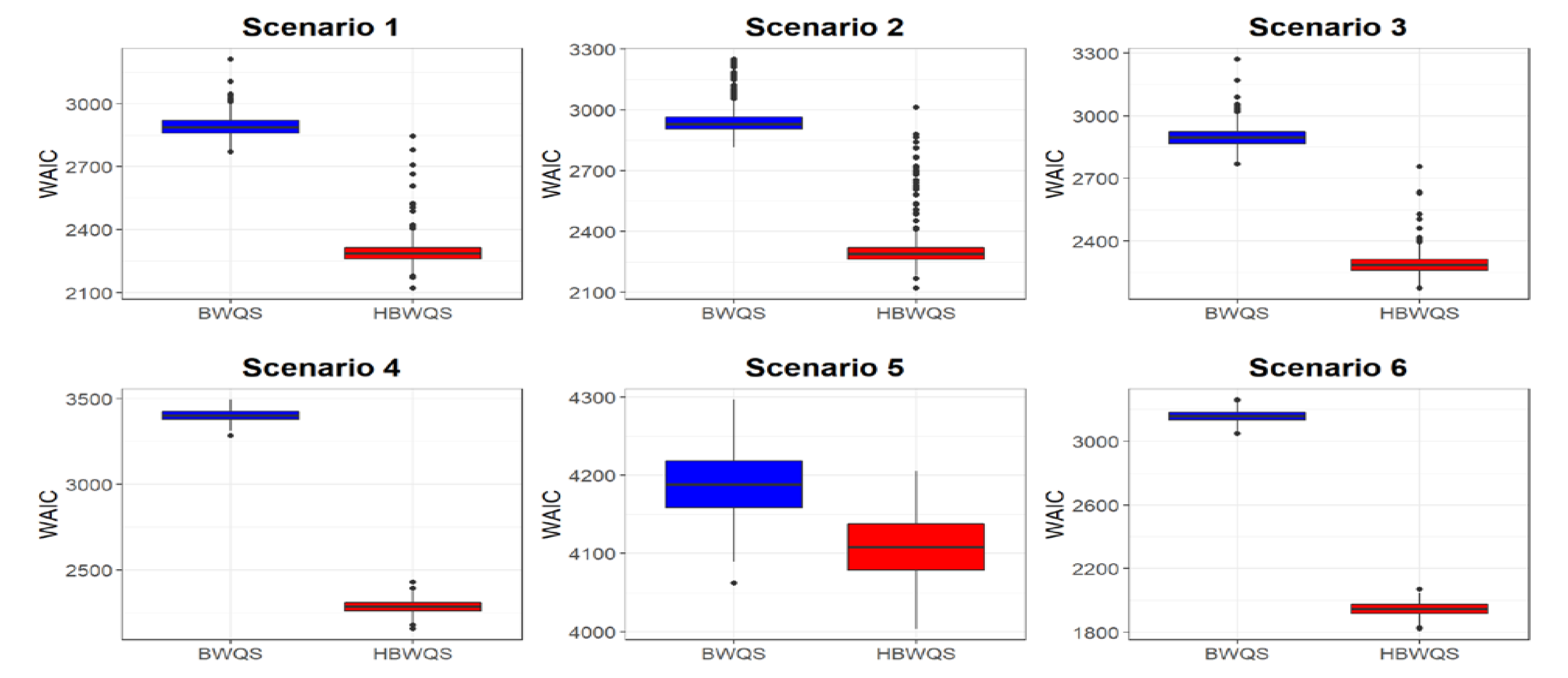
Watanabe-Akaike information criterion (WAIC) for the Bayesian Weighted Quantile Sum regression (BWQS) and the hierarchical BWQS (HBWQS) regression across six simulated scenarios.

Both HBWQS and BWQS models had reasonable computing time. The computing time for the HBWQS model required an average time of 7 minutes (426 seconds) for all the scenarios, except Scenario 5, which required 40 minutes (2,413 seconds), while the BWQS computing time required an average of 5 minutes (308 seconds) (**Table S3**). This is consistent with expectations since an increased model complexity and model variability imply more computing time to have an appropriate model convergence.

## 4. Real data applications: gestational age and gestational age acceleration in relation to metal mixture

### 4.1 Cohorts’ descriptions and ethics

#### The PRogramming of Intergenerational Stress Mechanisms (PRISM) study

The mother-infant pairs included in these analyses are participants in the PRISM prospective pregnancy cohort study designed to examine the effects of prenatal and early life psychosocial, physical, and chemical environmental exposures on child developmental outcomes. Beginning in 2011, pregnant women were recruited from prenatal clinics in Boston and New York City. Women were eligible if they were English or Spanish speaking, 18 years or older, and pregnant with a singleton. Exclusion criteria included maternal intake of ≥7 alcoholic drinks per week prior to pregnancy, any alcohol intake after pregnancy recognition, and HIV+ status. The study recruited n=1,109 women at 25 ± 7 weeks of gestation receiving prenatal care from the Beth Israel Deaconess Medical Center and East Boston Neighborhood Health Center in Boston, MA (from March 2011–December 2013) and Mount Sinai Hospital in New York City, NY (from April 2013– February 2020) who delivered a live newborn with no significant congenital anomalies noted at birth that could impact participation in follow-up procedures.^18, 19^ Metals and creatinine were measured in a subset who also had urine samples collected during pregnancy (N=641, 252 in Boston (BOS) and 389 in New York City (NYC)). Within two weeks of enrollment, mothers completed standardized surveys via in-person interviews to ascertain relevant covariates and exposures. Study protocols were approved by the Institutional Review Boards (IRBs) of the Brigham and Women’s Hospital and the Icahn School of Medicine at Mount Sinai. Mothers provided written consent in their primary language. In these analyses we split the PRISM study using the enrollment location, due to the differences in participants’ characteristics (**Table 1**) <u>The First 1000 Days and Beyond (F1000 or FTDL) study</u> is a longitudinal pregnancy cohort of mother-child pairs living in Virginia.^20, 21^ Women were ≥18 years of age and willing to participate in 12 longitudinal surveys during the first 12 months postpartum. All women were enrolled in 2012 at 26 ± 5 weeks’ gestation at the Inova Health System in Falls Church, VA for the genetics study. Starting in 2016, participants from the ongoing F1000 study were invited to participate in the national ECHO program, with 1,400 mothers consenting for themselves and their children to participate between 2016 and 2021.^22, 23^ A total of 708 mother-child pairs with complete data on birth information also consented for biospecimen collection including urine for metal assays. The human studies committee at the George Mason University ceded review to the Mount Sinai IRB for ECHO follow-up and mothers provided written informed consent.

### 4.2 Primary birth outcomes

Gestational age and gestational age acceleration metrics are good indicators of the fetal environment and predictors of neonatal short-and long-term child health^24–26^

#### Gestational age

In PRISM, gestational age was calculated based on maternal report of last menstrual period (LMP) and updated, if discrepant by more than three weeks, based on obstetric estimates from ultrasound data in the medical record review at delivery.^27^ In F1000, ultrasounds were not performed routinely as standard of care; thus, gestational age was based on LMP and a standardized physical examination at birth to determine gestational age.^28, 29^

#### Gestational age acceleration metrics

In PRISM, we estimated three placental epigenetic gestational age estimators: a “robust placental clock” (RPC), a “control placental clock” (CPC) and a “refined RPC” (refRPC). ^30^ The RPC metric was developed to be unaffected by pregnancy conditions, such as preeclampsia, gestational diabetes, and trisomy; the CPC metric was designed for measuring gestational age in normal pregnancies, and the refRPC was trained for uncomplicated term (gestational age > 36 weeks) pregnancies.^30^ We then used the epigenetic gestational age estimators to compute gestation age acceleration metrics that were formally defined as studentized residuals resulting from regressing the gestational age estimators on observed gestational age. By definition, these residual-based measures of gestational age acceleration were not correlated with true gestational age (r=0).^30^ The three placental epigenetic gestational age metrics leveraged PRISM placenta DNA methylation data collected immediately following birth as previously described.^31, 32^ Briefly, sufficient DNA was available from 234 placenta samples and DNA methylation was measured using the 450K array (Illumina, San Diego, CA). To minimize batch effects, samples were randomized into chips and plates.^33, 34^ Raw methylation data were processed using the R package *ewastools.*^35, 36^ We removed samples that were outliers and were potentially contaminated, and we corrected for dye bias using RELIC.^37^ Observations with detection *p*-values < 0.05 were also removed.^35, 36^ In F1000, placenta DNA methylation data were not available, so analyses on gestational age acceleration metrics included only sites from the PRISM study.

### 4.3 Prenatal chemical exposures: Arsenic, Cadmium and Lead concentrations

In both cohorts, maternal urine collected during gestation was stored at −80°C, and shipped to Mount Sinai for laboratory analysis as previously described. ^19^ Urine samples (200 µl) were diluted with solution in a polypropylene trace-metal-free Falcon tube.^19^ Samples were analyzed using matrix-matched calibration standards using an 8800 triple-quad inductively coupled plasma tandem mass spectrometer (Agilent Technologies) with appropriate gases to eliminate ion interference. Internal standards were used to correct for differences in sample introduction, ionization, and reaction rates.^19^ To monitor the accuracy, recovery rates, and reproducibility of the procedure, quality control and quality assurance procedures included analyses of initial calibration verification standards and continuous calibration verification standards.^19^ The limits of detection for analytes by this procedure were between 0.02 and 10 ng/ml.^19^ When we had two urinary metal measurements during gestation, we considered the average exposure between the 2^nd^ and the 3^rd^ trimesters to reflect the entire pregnancy exposures.

### 4.4 Covariates

Both PRISM and F1000 studies included maternal age, race/ethnicity, parity, maternal education and child sex.^20, 26^ Urinary creatinine levels, available for both cohorts, were measured using a well-established colorimetric method (limit of detection [LOD]: 0.3125 mg/dL)^38^ and were used to correct metal levels for urinary dilution variation. Creatinine levels were averaged when we considered the simple average exposure between the 2^nd^ and the 3^rd^ trimesters. Individuals with missing covariate information or smoking during pregnancy, for consistency across studies and due to the strong smoke effect on birth outcomes, were removed from the analyses, as described in the flowchart (**Figure S3**).

### 4.5 Models

We log2-transformed all metals to reduce the skewness of their distributions and we harmonized the covariates to accommodate any major difference. We then employed the HBWQS regression to evaluate the association of each outcome with the urinary metal mixture including all cohorts. We also applied the BWQS regression in each cohort separately and combining all cohorts in a single analysis. All HBWQS models included a single chain of 20,000 iterations, whose 10,000 were part of the burn-in and we used 10 as thinning parameter, while all BWQS models had a single chain of 10,000 with 5,000 warm-up and 3 thin. We reported the quality of the convergence of the chain of each parameter using 1) trace and 2) autocorrelation plots, 3) Rhat coefficients, and 4) the number of effective sample size (ESS), which reflects the autocorrelation within the chain.^39, 40^ For the gestational age analyses, we first analyzed all n=1,349 samples (BOS n=252, NYC n=389, F1000 n=708, **Table 1**, **Figure S3**) and we then restricted our sample size to children born at term (gestational age ≥ 37 weeks) to mitigate any potential skewness of the outcome distribution, thus leaving n=1,276 (BOS n=238, NYC n=346, F1000 n=692) for the analyses. For the analyses on gestational age acceleration metrics, we considered only PRISM data, thus leaving n=170 (BOS n=106, NYC n=60, **Table 1**, **Figure S3**). We finally compared HBWQS and BWQS regression on the same data using the WAIC metrics and provided computing time for all analyses.

### 4.5 Results

The three studies showed differences across participants’ characteristics. Women living in NYC showed shorter gestational age (38.5 weeks) compared to the other studies (Gestational age: BOS: 39.2, F1000: 39.3, pvalue<0.001, **Table 1**) and delivered at younger age (28.1 years) compared to other locations (BOS: 32.0, F1000: 32.5, pvalue<0.001). We identified differences of race/ethnicity distribution across studies (pvalue<0.001), with higher proportion of Black (57%) and non-Black Hispanic (38%) in PRISM NYC, compared to PRISM BOS (Black: 21%, Non-Black Hispanic: 33%) and F1000 (Black: 4%, Non-Black Hispanic: 14%). No difference of gestational age acceleration was identified between PRISM BOS and PRISM NYC (**Table 1**). Despite the difference in mean metal concentrations across cohorts (pvalue<0.001; As PRISM BOS: 3.45 PRISM NYC: 3.42 F1000: 2.57; Cd: PRISM BOS:-2.83 PRISM NYC:-2.20 F1000:-3.04; Pb: PRISM BOS:-1.43 PRISM NYC:-0.65 F1000:-1.65, **Table 1**), the metals had common domains (**Figure 2A**) and similar correlation pattern (**Figure 2B**). To create a common scale, the metal domains across studies were used to compute the exposure quantiles. The correlation between metals was moderate in each cohort, ranging between 0.13-0.28 in PRISM BOS, between 0.18-0.24 in PRISM NYC and between 0.15-0.31 in F1000. All models showed a good convergence of the chains: 1) all trace plots show good evolution of the parameter vector over the Markov chain iterations (**Figures S4**-**S5**), 2) the autocorrelation across post-warmup iterations of the posterior distributions dropped quickly to zero with increasing lags, indicating no autocorrelation across iterations (**Figures S6**-**S7**), 3) Rhat statistics were approximately 1 for all parameters, indicating that the chain has converged to the equilibrium distribution (**Tables S4**-**S5**) and 4) the ratio between the effective sample size (ESS) and the total sample size of chain iterations of the posterior distributions was above 0.75 for all parameters, indicating good ability of the chain to estimate the true mean value of the parameters (**Figures S8**-**S9**, **Tables S4**-**S5**). We reported the computing time for the HBWQS and BWQS models on the same data (**Table S6**). As we showed in the simulated scenarios, more complex models, i.e. HBWQS regressions, had a higher computing time with than simple ones, i.e BWQS regressions.

**Figure 2.**
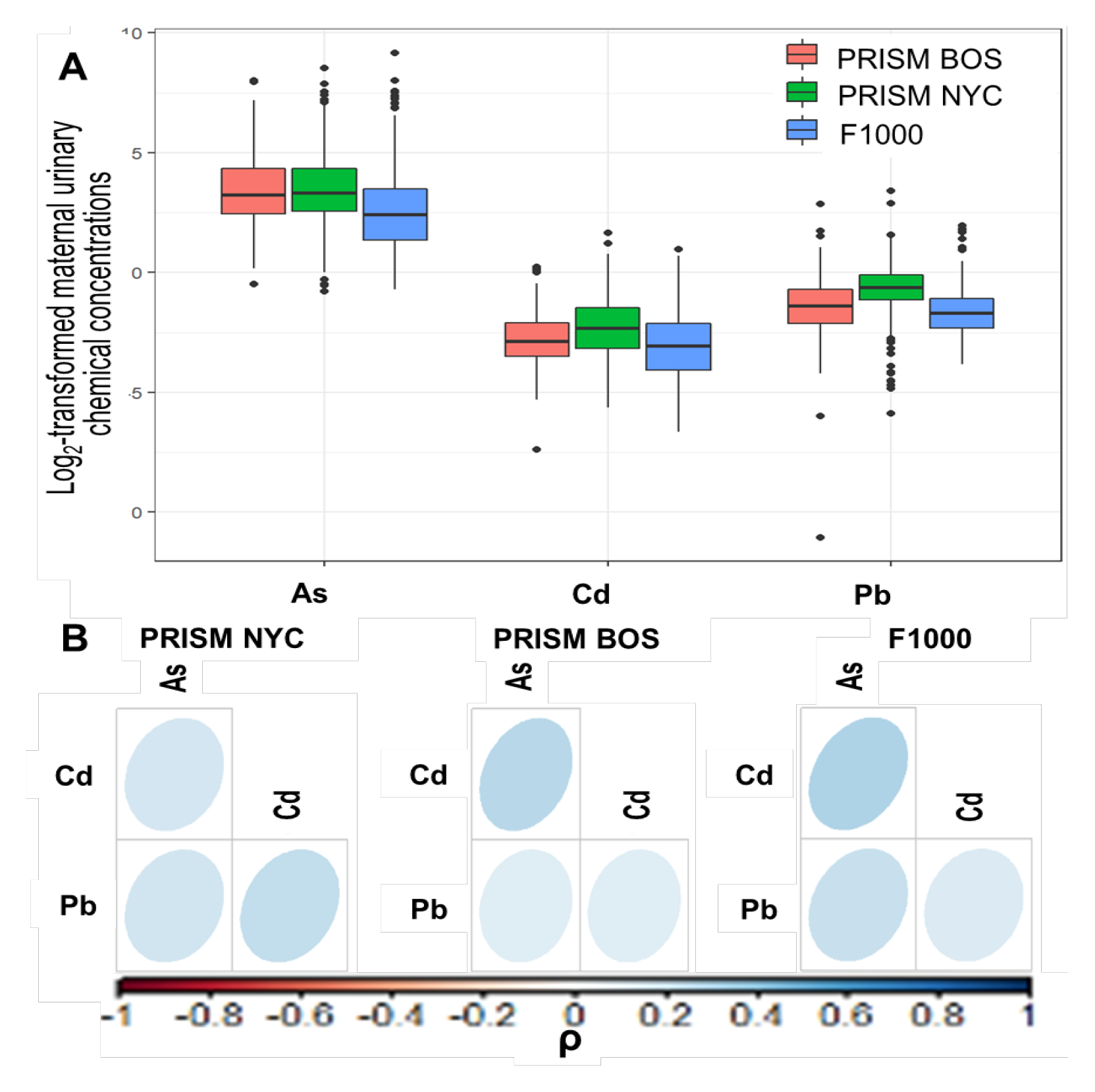
**A.** Boxplots of the log-2 transformed urinary metal levels (Arsenic (As), Cadmium (Cd), Lead, (Pb), in the three studies (PRISM Boston (BOS), PRISM New York City (NYC), F1000). **B** Pearson correlation plots of the log-2 transformed urinary metal levels within each study.

#### Gestational age

The HBWQS regression showed a negative association between the metal mixture and gestational age in the PRISM NYC (Est. −0.22; 95%CrI:-0.40; −0.02; 80%CrI: −0.34; −0.10, **Table S4**, **Figure 3**), with Cd (64%) contributing the most to the overall mixture effect (As: 21%; Pb:15, **Figure 3**). While using the BWQS regression, no evidence of association was identified between gestational age and metal mixture at any site separately (PRISM BOS: Est. 0.02; 95%CrI:-0.23; 0.28; 80%CrI: −0.15; 0.19; PRISM NYC: Est. −0.11; 95%CrI:-0.53; 0.32; 80%CrI: −0.40; 0.17; F1000: Est. −0.05; 95%CrI:-0.18; 0.08; 80%CrI: −0.13; 0.03 **Table S4**, **Figure 3**) or in combination (Est. −0.09; 95%CrI:-0.24; 0.06; 80%CrI: −0.19; 0.01 **Table S4**, **Figure 3**). The HBWQS showed a better performance (WAIC: 5180) than the BWQS regression on the combined sites (WAIC: 5200) (95%CrI of the WAIC difference: 1.00; 39.80). Once we included only women that delivered at term (≥37 weeks) (n=1,276), the majority of results were consistent with previous analyses showing no association between the metal mixture and gestational age in individual sites or in combination. Although HBWQS findings showed no longer a significant association between gestational age and the metal mixture in PRISM NYC, the relationship was consistent in the directionality (**Table S4**, **Figure S10**).

**Figure 3.**
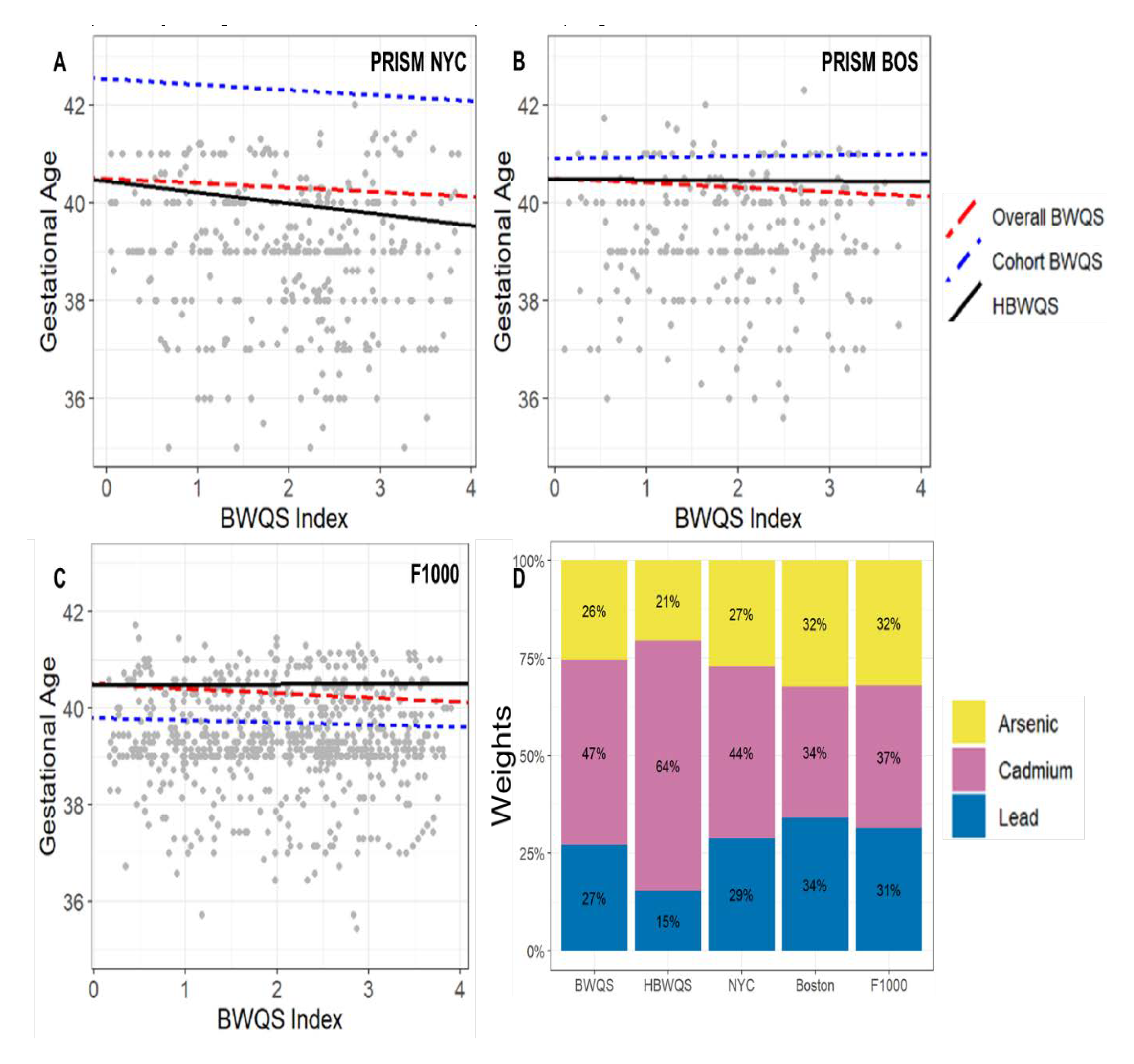
**A-C** Estimated association between the prenatal urinary metal mixture exposure (Arsenic (As), Cadmium (Cd), and Lead (Pb)) and gestational age (weeks) in the PRISM New York City (NYC), PRISM Boston (BOS) and F1000 by using the Bayesian weighted quantile (BWQS) regression at the individual sites (cohort BWQS) and in the overall sample aggregating all sites (Overall BWQS) and by using the hierarchical BWQS (HBWQS) regression. **D** Contribution of metals to the mixture.

#### Gestational age acceleration

There was no evidence of the association between the metal mixture and the gestational age acceleration metrics in the overall PRISM study combining both sites with BWQS regressions (RPC metric: Est. −0.05; 80%CrI: −0.18; 0.08; CPC metric: Est. - 0.3 80%CrI: −0.16,0.10; refRPC metric: Est. −0.10; 80%CrI: −0.22; 0.03 **Table S5**, **Figure 4**) or using the HBWQS approach (RPC metric: PRISM BOS: Est. −0.02; 80%CrI: −0.15; 0.12; PRISM NYC: Est. −0.08; 80%CrI: −0.24; 0.06; CPC metric: PRISM BOS: Est. 0.00; 80%CrI: −0.14; 0.14; PRISM NYC: Est. −0.05; 80%CrI: −0.20; 0.10; refRPC metric: PRISM BOS: Est. −0.10; 80%CrI: - 0.24; 0.04; PRISM NYC: Est. −0.13; 80%CrI: −0.29; 0.03 **Table S5**, **Figure 4**). However, all estimates showed consistent negative directionality and similar magnitude of the mixture effects on the outcomes. Across gestational age acceleration metrics, HBWQS WAIC values were similar to those of the BWQS model combining the two sites (RPC metric: HBWQS: 461 BWQS: 460; 95%CrI WAIC difference: −3.8; 1.7; CPC: HBWQS: 461 BWQS: 461; 95%CrI WAIC difference: −3.3; 3.0 refRPC: HBWQS: 462 BWQS: 460; 95%CrI WAIC difference: −3.1; 0.1) and under these circumstances the simplest BWQS analysis is preferable than the HBWQS regression with random terms for site. We identified a weak evidence of a negative association between the metal mixture and both RPC (Est. −0.34; 95%CrI:-0.78; 0.10; 80%CrI: −0.63; −0.05), and refRPC (Est. −0.31; 95%CrI:-0.74; 0.12; 80%CrI: −0.59; −0.03) in the PRISM NYC study, probably due to the higher homogeneity of this population. In both associations, Pb (RPC: 46%, refRPC: 41%) and Cd (RPC: 34%. refRPC: 37%) showed the highest contribution to the mixture (**Table S5**, **Figure 4A-B**).

**Figure 4.**
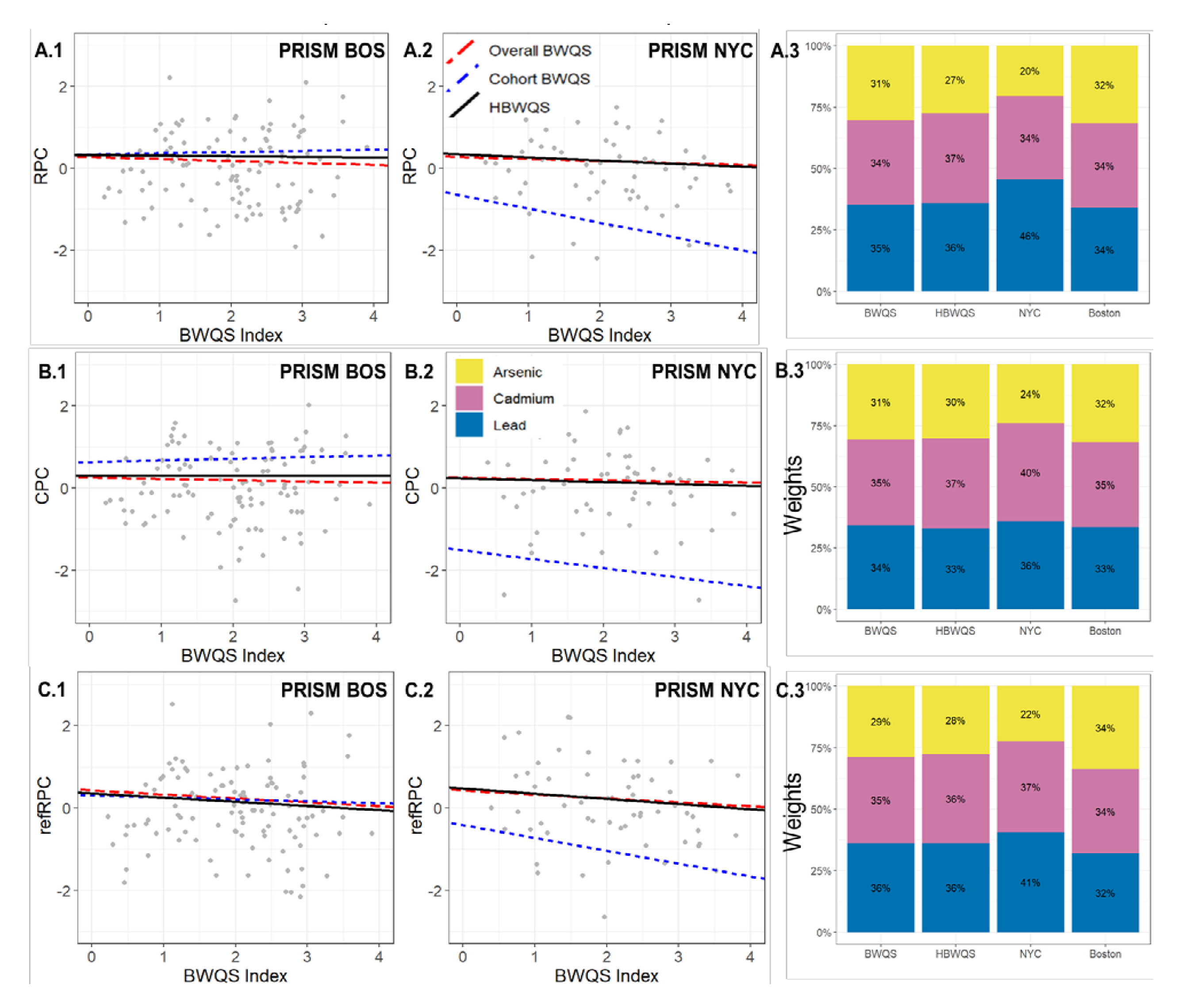
**A.1-A.2, B1-B.2, C.1-C2** Estimated association between the prenatal urinary metal mixture exposure (Arsenic (As), Cadmium (Cd), and Lead (Pb)) and gestational age acceleration metrics (RPC, CPC, refRPC) in the PRISM New York City (NYC), and PRISM Boston (BOS) by using the Bayesian weighted quantile (BWQS) regression at the individual sites (cohort BWQS) and in the overall PRISM sample aggregating all sites (Overall BWQS) and by using the hierarchical BWQS (HBWQS) regression. **A.3, B.3, C.3** Contribution of metals to the mixture. RPC= robust placental clock; CPC: control placental clock; refRPC: refined RPC

## 5. Discussion

We developed and applied the HBWQS regression, which estimates (i) an overall mixture index for the exposures and (ii) cohort-specific regression coefficients that characterize the association between an outcome and this overall mixture index. The overall mixture is a weighted index that linearly combines the exposures that can be previously ranked in quantiles. The mixture weights are assumed to follow a Dirichlet distribution, thus implying that each weight is constrained to be positive and the sum of all weight is constrained to be 1, so that they can be sorted by relative importance. Exposures that affect the outcome to a greater degree have higher weights, whereas exposures with little or no impact on the outcome have near-zero weights. The HBWQS method uses random slopes to infer the association between the outcome and the overall weighted mixture index in each cohort. Our simulations showed that the parameters estimated with HBWQS regression showed very little bias and had a good empirical coverage across scenarios. The MSEs were small and close to zero for the majority of parameters. We also compared the HBWQS and BWQS regressions using the WAIC and simulation findings showed that the HBWQS performed better than BWQS regressions on the same datasets.

This approach offers several advantages. First the overall mixture index is based on the quantile for each metal exposure, thereby reducing the impact of extreme values in right-skewed distributions. This step also creates a common scale across cohorts for each exposure, because the quantiles are created using the exposure data from all cohorts^15^. Second, the weights w_j_ are constrained to be positive and sum to 1 so that all exposures in the mixture are assessed simultaneously and incorporated into a single index.^15^ These weights give the relative importance of the exposures, identifying which exposure within the mixture is the most relevant into the mixture.^15^ Third, the hierarchical form of this approach and specifically the random effect terms enable us to have a cohort-specific association between the outcome and the overall mixture index and a cohort-specific intercept, respectively. In addition, this hierarchical method incorporates the differences in exposures correlation structure of each cohort with random effects and facilitates sharing information across cohorts as all hierarchical Bayesian approaches do. ^41^ The advantages of information sharing of hierarchical modeling include the shrinkage of extreme coefficient estimates towards an overall average and the fewer observations a cluster, i.e. cohort, has the more information is borrowed from other clusters and the greater the pull towards the average estimate is.^42, 43^ Finally, HBWQS regression capitalize upon the idea that independent studies are randomly sampled from a population of studies, and individual subjects are randomly sampled within each study.^41^ The combination of multiple individual studies, which are thought to be sub-studies from a population of studies, substantially increases the statistical power for testing research hypothesis and integrates between-study heterogeneity. Indeed, when multiple independent studies are combined, there is often an increase in statistical power when testing the same hypotheses based upon the aggregated versus independent studies.^44^ Also the between-study heterogeneity may serve to have more generalizable conclusions on the composition of the overall mixture index and may offer novel insights of mixture-outcome associations in each individual study. This approach may help mitigate the need for creating novel studies designed to resolve conflicting findings across studies posited to result from between-study design differences.

We applied the HBWQS regression to three ethnically diverse populations sharing the same outcomes, exposures (As, Cd, and Pb) and harmonized covariates. We found a detrimental negative association between the metal mixture and gestational age in the PRISM NYC sample using the HBWQS regression. The mixture was mostly composed by Cd (64%). Although there was no evidence of the mixture associations with gestational age acceleration metrics using HBWQS and BWQS on aggregated sites, all results suggested a negative direction of the associations. We also uncovered a weak evidence of a negative association between the metal mixture and gestational age acceleration (both RPC and refRPC) in the PRISM NYC sample using BWQS analysis with Cd and Pb having higher contribution to the mixture. This set of results suggested a slower biological gestational age with higher exposure in the metal mixture.

Our results are consistent with prior literature showing prenatal individual and mixture exposure to arsenic (As), lead (Pb), and cadmium (Cd) associated with shorter gestation ^45–48^, and preterm birth ^47, 49–51^ in populations exposed to chemicals at different levels of exposure. However, results considering environmental mixtures are still conflicting on showing the most detrimental exposures into the mixture and the significance of the overall the mixture-outcome associations.^48, 52–55^ Overall, our application highlighted the importance of evaluating metal mixture in relation to children health, including birth outcomes. Indeed, compared to adults, children have differences in metal exposure patterns and have greater vulnerability to metal exposures due to crawling, hand-mouth behaviors, and diet.^56^ In addition, environmental metal exposures may be higher than adults due to higher surface-to-volume ratios, higher consumption rates of food and water, higher respiration rates, and different toxicokinetics.^56^ Although high levels of many individual metal exposures, such as lead, produce observable toxicity early in life, including on gestational age, lower doses of multiple metals may be detrimental.^57^ During critical developmental periods, including gestation, low levels of numerous metal exposures might induce only subtle alterations in endocrine signaling or gene expression but could lead to permanent changes that program health outcomes throughout the life course.^57^

Our real case studies supported the idea that analyses at individual heterogeneous studies may be limited by sample size to capture small effect sizes, which can be different across studies and cannot be identified in aggregated analyses. This novel approach, however, cannot be blindly used for combining data from any independent studies, and some considerations should be assessed before applying the HWQBS regression. Although study harmonization is the first step for developing commensurate measures and a good HBWQS application, it may be not sufficient. Indeed, even identical variables which do not require harmonization may function differently across studies, due to differences in regional interpretations, historical periods, and laboratory conditions and technologies. ^58^ For example, the same construct may not have equivalent meaning across studies of children at different ages.^44^ Also, even when the same study-characteristics are assessed with the same instrument in different laboratories, some technical artifacts may lead to some discrepancies across studies. ^44^

In addition, differences in study design characteristics may limit the applicability of this approach. For example, cross-study discrepancies in exposure ranges or socio-demographic characteristics can largely increase the sources of between-study variability and thus limiting the applicability of this approach. ^44^ Also, information missing by design (e.g. race/ethnicity) in a single cohort may be challenging to address using this approach, which requires to constrain the analysis to common items. Although a missing at random assumption could seem quite plausible in many cases, the exclusion of the missing variable should be evaluated with sensitivity analyses and alternatives approaches, such as residual analysis. Also, different study-designs, such as case-control vs observational studies, may be not appropriately treated in a combined analysis with random effects and fixed-effects approach, i.e. BWQS regressions, should considered instead. ^44^ Finally there must be a sufficient number of independent samples and studies to allow for the reliable estimation of the random variability. Although further research can help to detect the sufficient number of samples and studies to allow for proper estimation of mixture-outcome association with random effects, prior general multilevel framework suggested between 20 to 30 as a minimum number of samples, while three as the minimum number of studies.^44, 59^ The influences of all these potential sources of between-study heterogeneity should be considered and mitigated when possible before applying HBWQS regression in order to optimally capture the estimates of the mixture-outcome associations and mixture component contribution.

Overall, the HBWQS approach directly supports the increased calls for data sharing, the maximization of available resources and the creation a new research endeavors, where researchers can contribute with secondary data analyses, and multiple innovative methodologies thus offering novel insights of the data. The analysis of extant data is an extremely cost efficient mechanism for conducting research and this efficiency is further realized by considering not just one but multiple existing datasets with same exposures and outcomes.^60^

## 6. Conclusions

The HBWQS regression is the first mixture approach that can be applied to multiple cohorts and that can illustrate the discrepancies in the mixture-outcome associations across cohorts and characterize the individual contribution of each component to the overall mixture index.

## Data Availability

For all data produced in the present study please contact the corresponding author

## ECHO Collaborators Acknowledgments

The authors wish to thank our ECHO Colleagues; the medical, nursing, and program staff; and the children and families participating in the ECHO cohorts. We also acknowledge the contribution of the following ECHO Program collaborators:

ECHO Components—Coordinating Center: Duke Clinical Research Institute, Durham, North Carolina: Smith PB, Newby LK; Data Analysis Center: Johns Hopkins University Bloomberg School of Public Health, Baltimore, Maryland: Jacobson LP; Research Triangle Institute, Durham, North Carolina: Catellier DJ; Person-Reported Outcomes Core: Northwestern University, Evanston, Illinois: Gershon R, Cella D.

ECHO Awardees and Cohorts—Boston Children’s Hospital, Boston, MA: Bosquet-Enlow M.

## Funding Acknowledgments and Disclaimer

The content is solely the responsibility of the authors and does not necessarily represent the official views of the National Institutes of Health.

Research reported in this publication was supported by the Environmental influences on Child Health Outcomes (ECHO) Program, Office of the Director, National Institutes of Health, under Award Numbers U2COD023375 (Coordinating Center), U24OD023382 (Data Analysis Center), U24OD023319 with co-funding from the Office of Behavioral and Social Science Research (PRO Core), 5U2COD023375-06 (Colicino), UH3OD023337 (Wright).

## SUPPLEMENTAL MATERIAL

**Figure S1.**
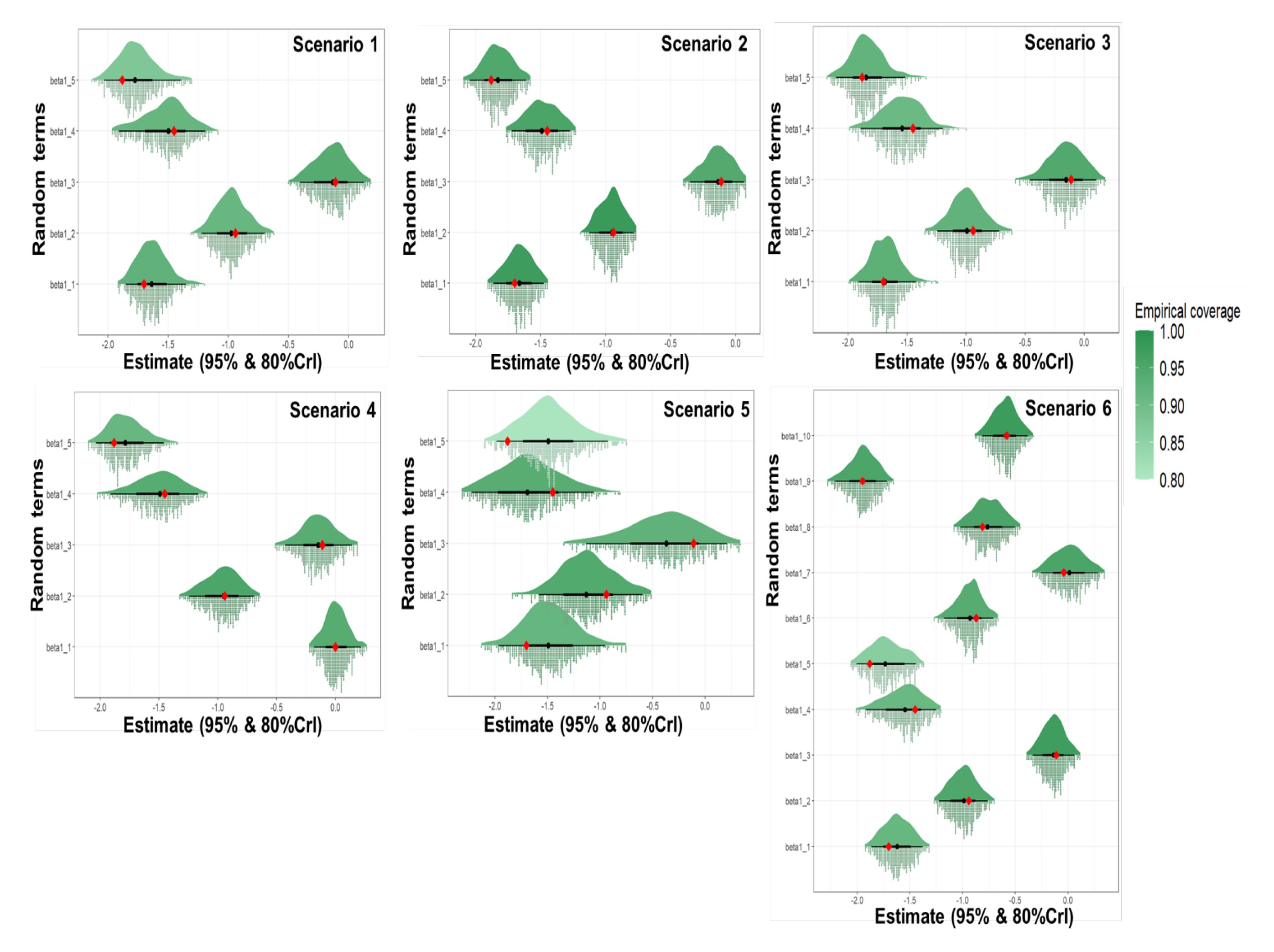
Real value (red dot), estimate (black dot), 95% and 80% Credible Interval (CrI) (thin and bold error bars), empirical coverage (color), and estimate distribution (distribution on the top, histogram on the bottom) of the random terms of the six simulated scenarios using the hierarchical Bayesian weighted quantile sum regression (HBWQS).

**Figure S2.**
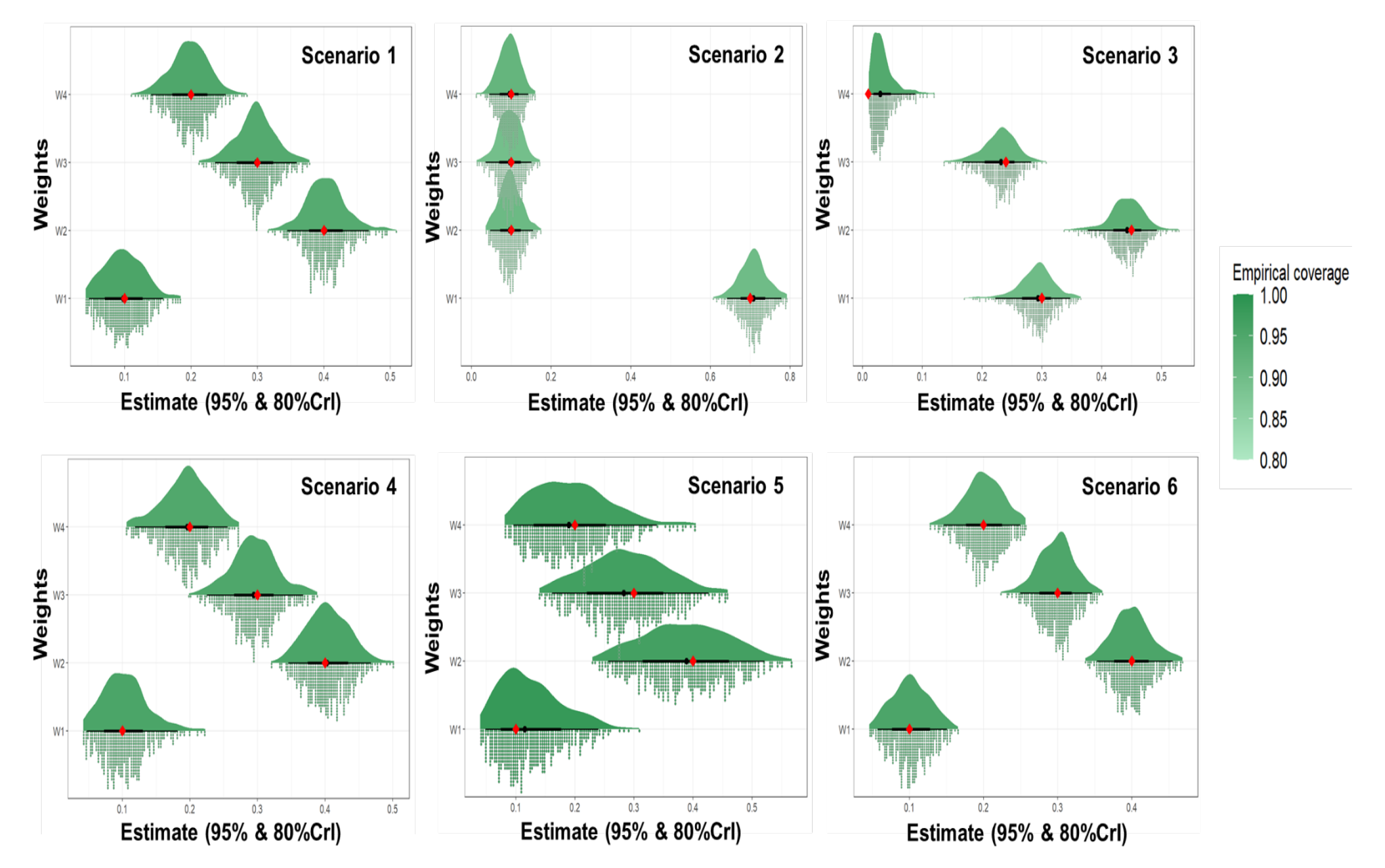
Real value (red dot), estimate (black dot), 95% and 80% Credible Interval (CrI) (thin and bold error bars), empirical coverage, and estimate distribution (distribution on the top, histogram on the bottom) of the weights of the six simulated scenarios using the hierarchical Bayesian weighted quantile sum regression (HBWQS).

**Figure S3.**
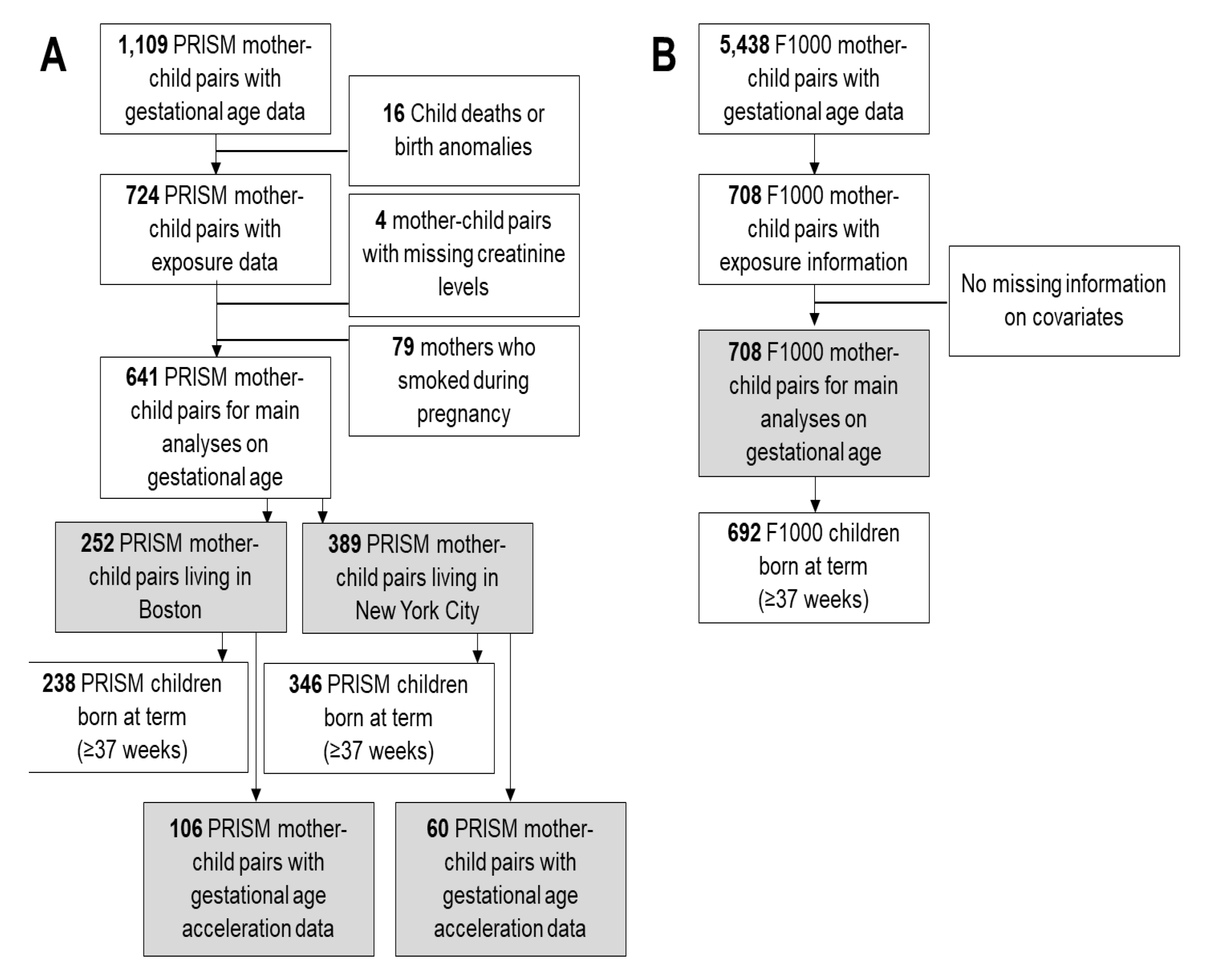
Flowchart for **A)** The PRogramming of Intergenerational Stress Mechanisms (PRISM) study and **B)** The First 1000 Days and Beyond (F1000) study. Grey boxes indicate the study subsets for the main analysis on both gestational age and gestational age acceleration metrics.

**Figure S4.**
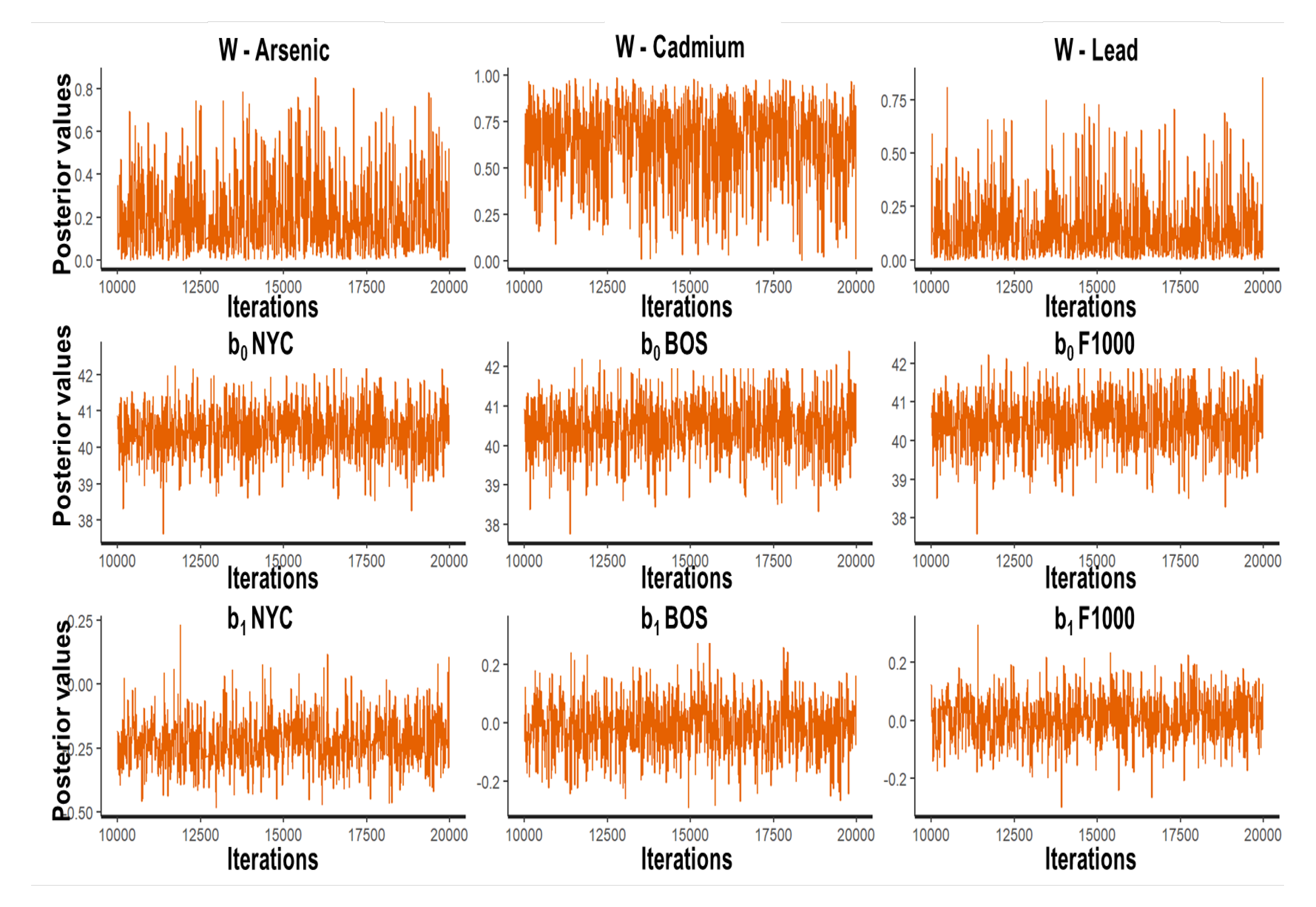
Trace plots of the post-warmup (10,000-20,000) iterations (X-axis) of the posterior distribution (Y-axis) of all main parameters (random intercepts: b_0_, random intercept: b_1_, weights: w)) of the hierarchical Bayesian weighted quantile (HBWQS) regression linking prenatal urinary metal mixture exposure (Arsenic (As), Cadmium (Cd), and Lead (Pb)) and gestational age (weeks) in the PRISM New York City (NYC), PRISM Boston (BOS) and The First 1000 Days and Beyond (F1000) studies.

**Figure S5.**
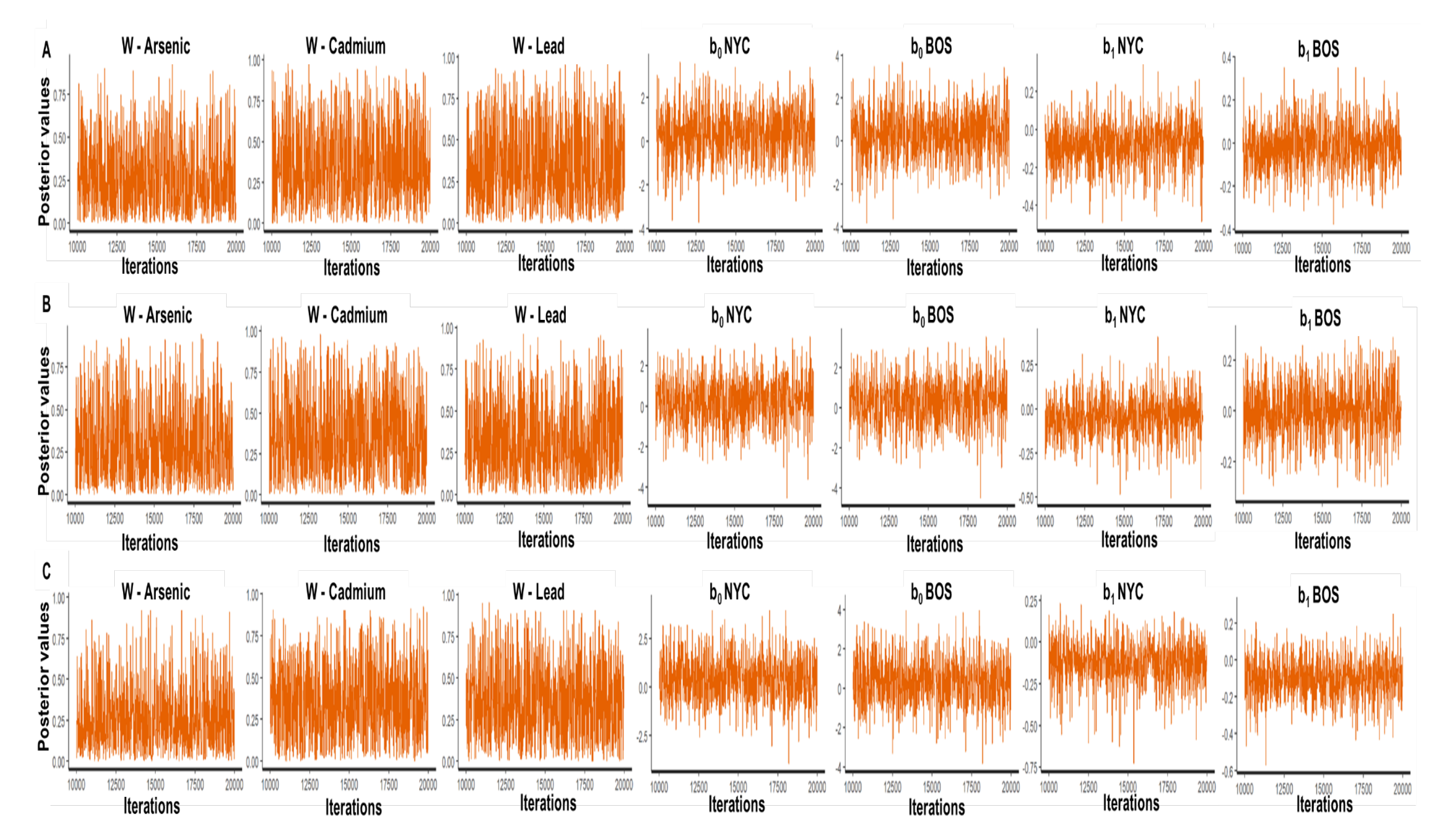
Trace plots of the post-warmup (10,000-20,000) iterations (X-axis) of the posterior distribution (Y-axis) of all main parameters (random intercepts: b_0_, random intercept: b_1_, weights: w)) of the hierarchical Bayesian weighted quantile (HBWQS) regression linking prenatal urinary metal mixture exposure (Arsenic (As), Cadmium (Cd), and Lead (Pb)) and gestational age acceleration metrics (**panel A.** RPC= robust placental clock; **panel B.** CPC: control placental clock; **panel C.** refRPC: refined RPC) in the PRISM New York City (NYC), and PRISM Boston (BOS).

**Figure S6.**
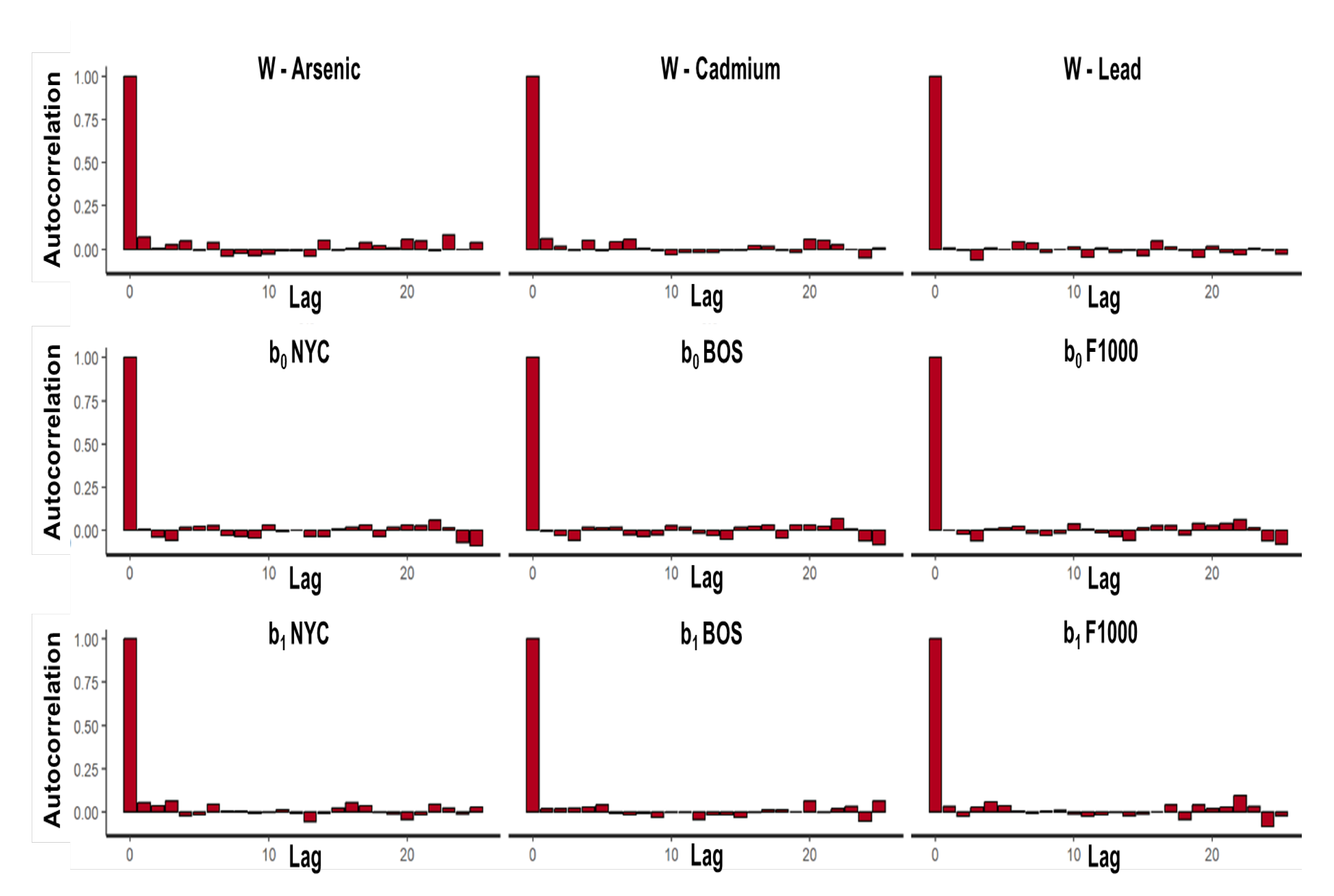
Autocorrelation plots of the Markov chain of the posterior distributions (Y-axis)—separately up to 25 lags (X-axis). We showed the posterior distributions of all main parameters (random intercepts: b_0_, random intercept: b_1_, weights: w)) of the hierarchical Bayesian weighted quantile (HBWQS) regression linking prenatal urinary metal mixture exposure (Arsenic (As), Cadmium (Cd), and Lead (Pb)) and gestational age (weeks) in the PRISM New York City (NYC), PRISM Boston (BOS) and The First 1000 Days and Beyond (F1000) studies.

**Figure S7.**
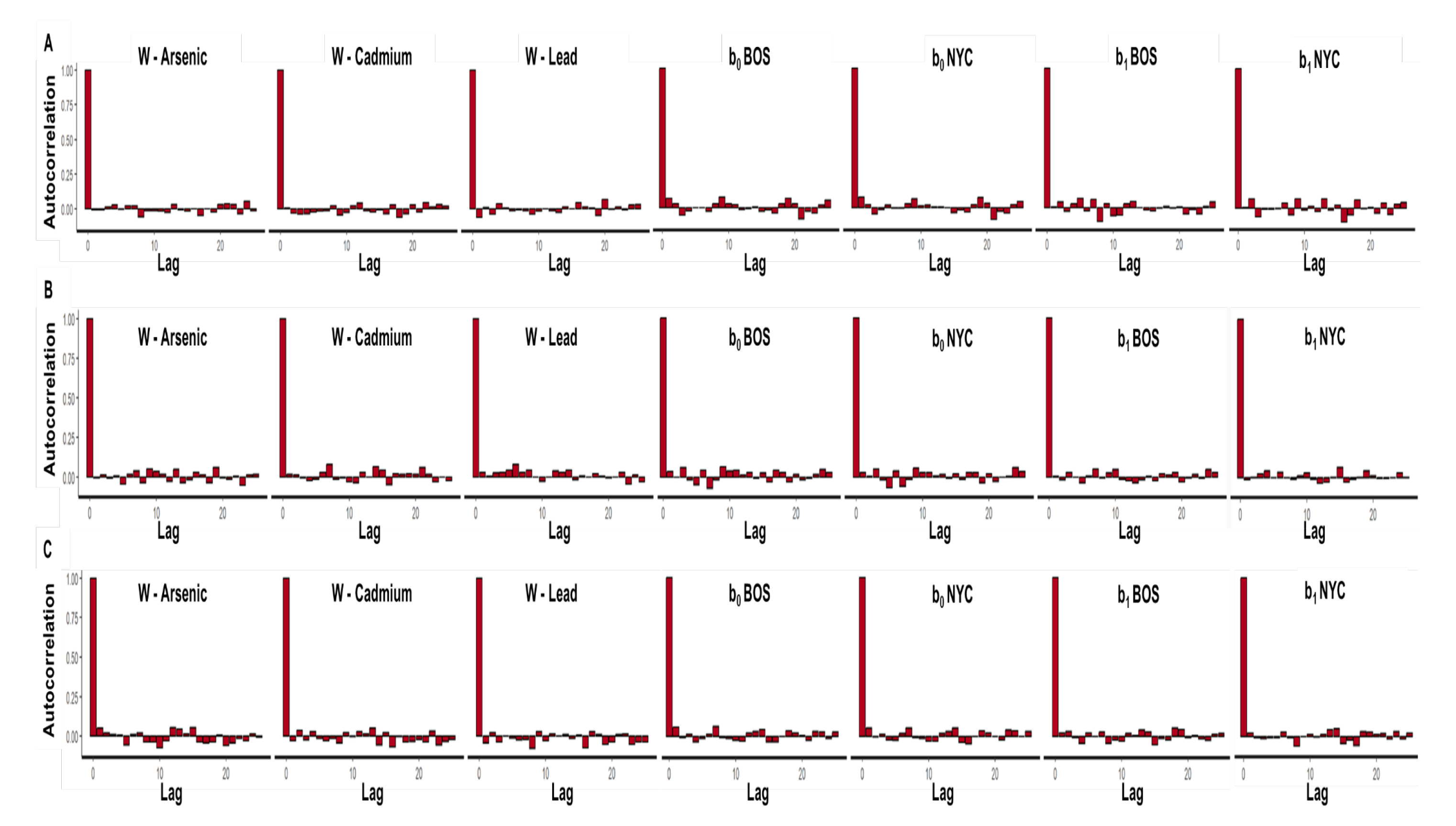
Autocorrelation plots of the Markov chain of the posterior distributions (Y-axis)—separately up to 25 lags (X-axis). We showed the posterior distributions of all main parameters (random intercepts: b_0_, random intercept: b_1_, weights: w)) of the hierarchical Bayesian weighted quantile (HBWQS) regression linking prenatal urinary metal mixture exposure (Arsenic (As), Cadmium (Cd), and Lead (Pb)) and and gestational age acceleration metrics (**panel A.** RPC= robust placental clock; **panel B.** CPC: control placental clock; **panel C.** refRPC: refined RPC) in the PRISM New York City (NYC), and PRISM Boston (BOS).

**Figure S8.**
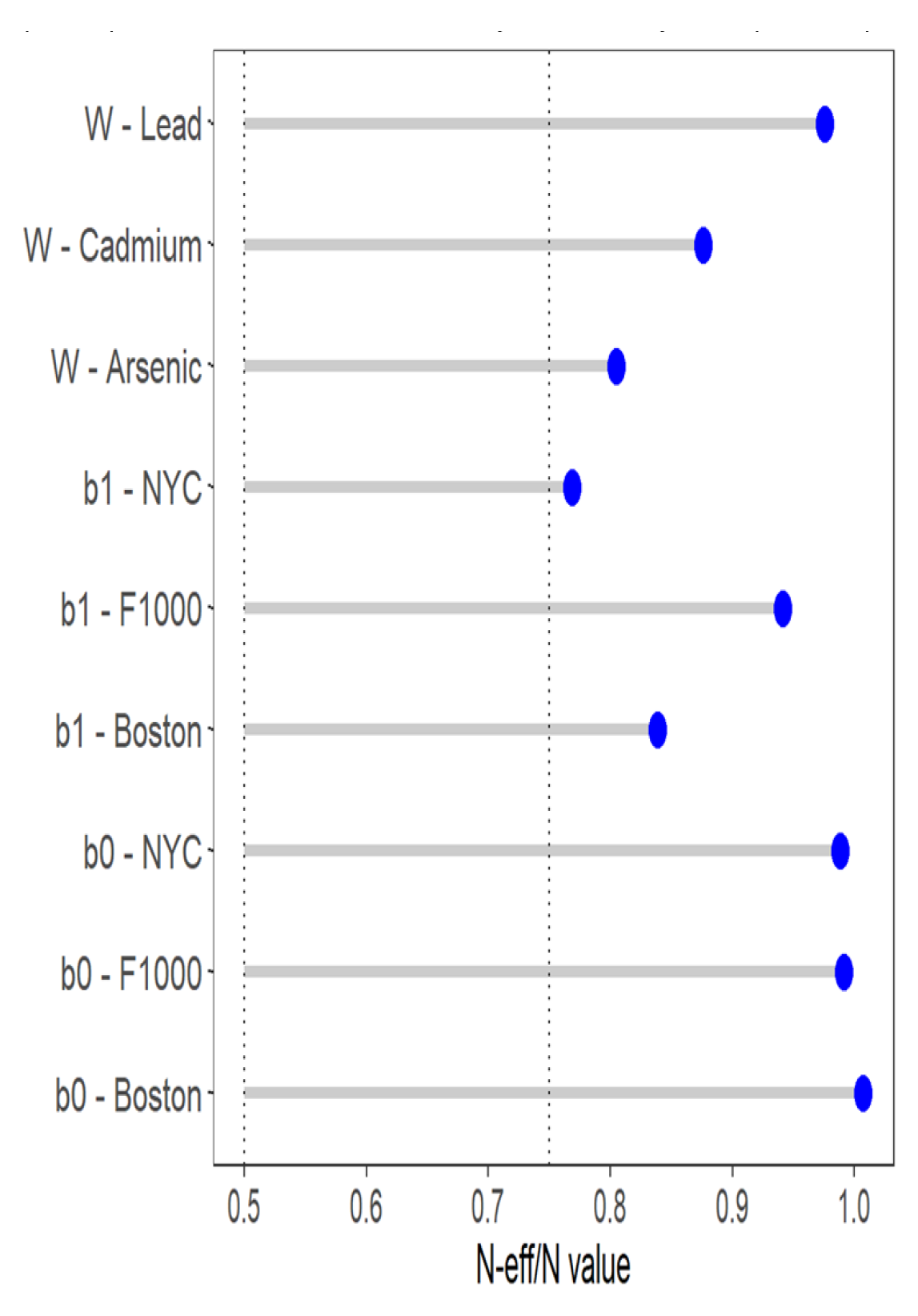
Ratio between the effective sample size (ESS) and total sample size of chain draws of the posterior distribution for main parameters(random intercepts: b_0_, random intercept: b_1_, weights: w)) of the hierarchical Bayesian weighted quantile (HBWQS) regression linking prenatal urinary metal mixture exposure (Arsenic (As), Cadmium (Cd), and Lead (Pb)) and gestational age (weeks) in the PRISM New York City (NYC), PRISM Boston (BOS) and The First 1000 Days and Beyond (F1000) studies.

**Figure S9.**
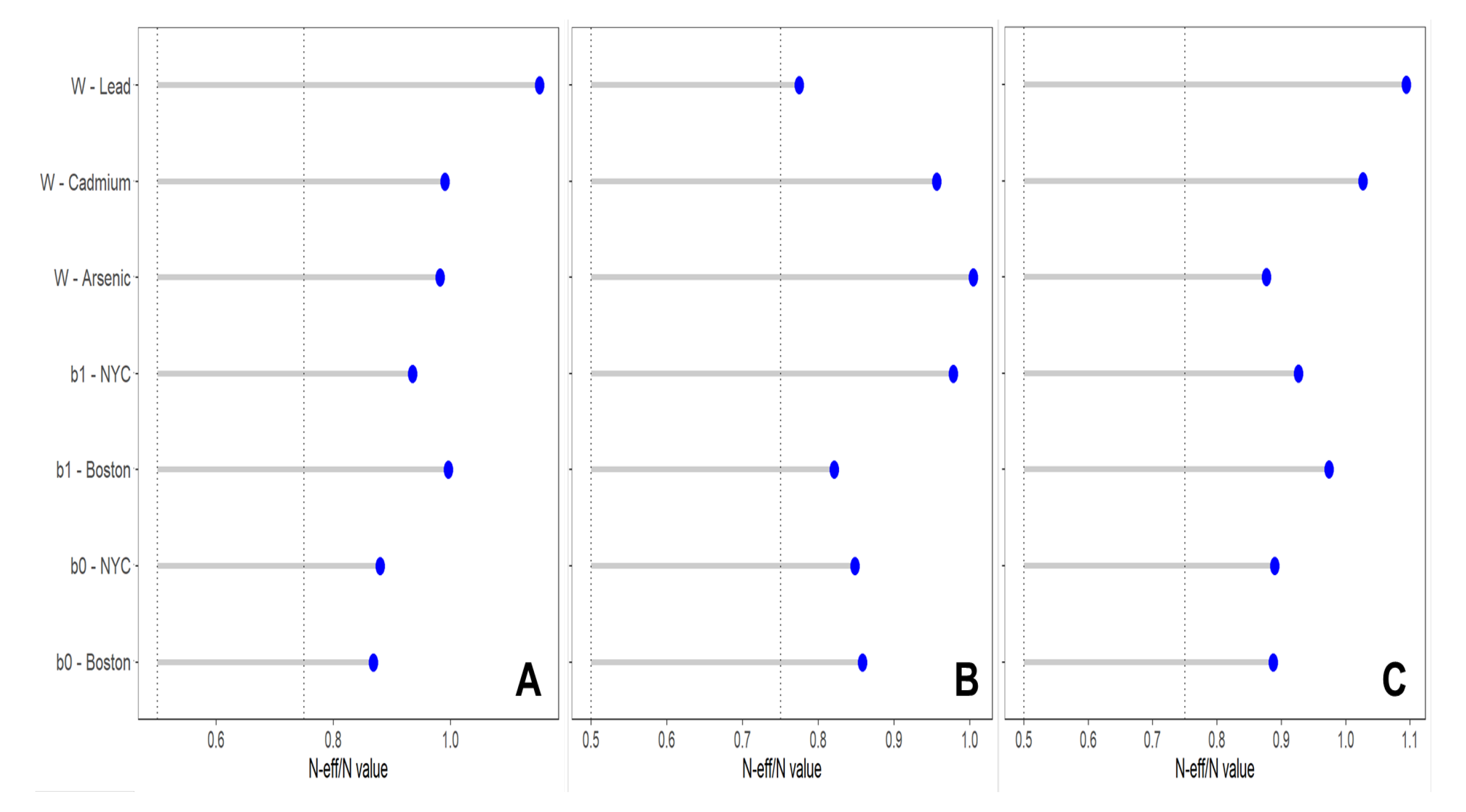
Ratio between the effective sample size (ESS) and total sample size of chain draws of the posterior distribution for main parameters(random intercepts: b_0_, random intercept: b_1_, weights: w)) of the hierarchical Bayesian weighted quantile (HBWQS) regression linking prenatal urinary metal mixture exposure (Arsenic (As), Cadmium (Cd), and Lead (Pb)) and and gestational age acceleration metrics (**panel A.** RPC= robust placental clock; **panel B.** CPC: control placental clock; **panel C.** refRPC: refined RPC) in the PRISM New York City (NYC), and PRISM Boston (BOS) studies.

**Figure S10.**
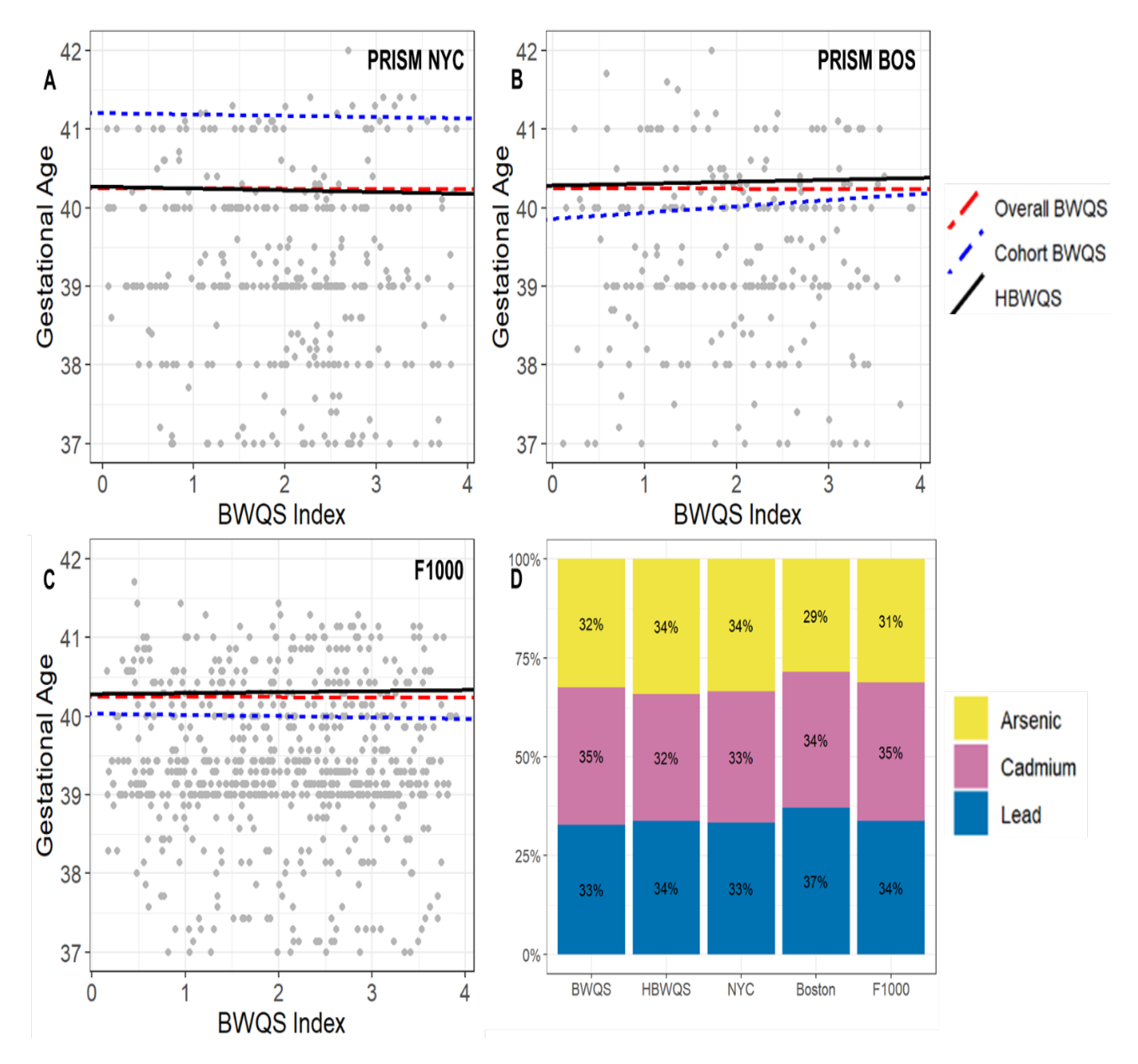
**Sensitivity analysis including only women that delivered at term (≥37 weeks) (n=1,276). A-C** Estimated association between the prenatal urinary metal mixture exposure (Arsenic (As), Cadmium (Cd), and Lead (Pb)) and gestational age (weeks) in the PRISM New York City (NYC), PRISM Boston (BOS) and F1000 by using the Bayesian weighted quantile (BWQS) regression at the individual sites (cohort BWQS) and in the overall sample aggregating all sites (Overall BWQS) and by using the hierarchical BWQS (HBWQS) regression. **D** Contribution of metals to the mixture.

**Table S1.** Real value, mean estimate, bias, Mean Square Error (MSE) and empirical coverage of the main parameters (random intercepts: beta_0_, random intercept: beta_1_, weights: W, model variance: sigma) of the six simulated scenarios using the hierarchical Bayesian weighted quantile sum regression (HBWQS).

| Parameters | Real value | Mean estimate | Bias | MSE | Empirical Coverage |
| --- | --- | --- | --- | --- | --- |
| Simulation 1 |  |  |  |  |  |
| beta0_1 | 0.65 | 0.45 | -0.20 | 0.55 | 0.93 |
| beta0_2 | -0.38 | -0.32 | 0.06 | 0.32 | 0.92 |
| beta0_3 | 0.15 | 0.24 | 0.09 | 0.23 | 0.95 |
| beta0_4 | -0.98 | -0.87 | 0.11 | 0.56 | 0.92 |
| beta0_5 | 1.13 | 0.83 | -0.30 | 0.69 | 0.90 |
| beta1_1 | -1.70 | -1.60 | 0.10 | 0.11 | 0.92 |
| beta1_2 | -0.94 | -0.97 | -0.03 | 0.05 | 0.91 |
| beta1_3 | -0.11 | -0.16 | -0.05 | 0.03 | 0.94 |
| beta1_4 | -1.45 | -1.50 | -0.05 | 0.11 | 0.92 |
| beta1_5 | -1.88 | -1.74 | 0.14 | 0.14 | 0.88 |
| W1 | 0.10 | 0.11 | 0.01 | 0.01 | 0.96 |
| W2 | 0.40 | 0.40 | 0.00 | 0.00 | 0.94 |
| W3 | 0.30 | 0.30 | 0.00 | 0.00 | 0.94 |
| W4 | 0.20 | 0.20 | 0.00 | 0.00 | 0.95 |
| Covariate1 | -0.40 | -0.40 | 0.00 | 0.00 | 0.94 |
| Covariate2 | 0.50 | 0.50 | 0.00 | 0.01 | 0.96 |
| Covariate3 | 0.20 | 0.20 | 0.00 | 0.00 | 0.96 |
| sigma | 1.00 | 1.00 | 0.00 | 0.58 | 0.94 |
| Simulation 2 |  |  |  |  |  |
| beta0_1 | 0.65 | 0.60 | -0.05 | 0.15 | 0.95 |
| beta0_2 | -0.38 | -0.32 | 0.06 | 0.14 | 0.95 |
| beta0_3 | 0.15 | 0.22 | 0.07 | 0.16 | 0.95 |
| beta0_4 | -0.98 | -0.87 | 0.11 | 0.17 | 0.97 |
| beta0_5 | 1.13 | 1.03 | -0.10 | 0.18 | 0.95 |
| beta1_1 | -1.70 | -1.67 | 0.03 | 0.01 | 0.97 |
| beta1_2 | -0.94 | -0.96 | -0.02 | 0.01 | 0.98 |
| beta1_3 | -0.11 | -0.14 | -0.03 | 0.02 | 0.95 |
| beta1_4 | -1.45 | -1.49 | -0.04 | 0.02 | 0.96 |
| beta1_5 | -1.88 | -1.83 | 0.05 | 0.02 | 0.95 |
| W1 | 0.70 | 0.71 | 0.01 | 0.00 | 0.97 |
| W2 | 0.10 | 0.10 | 0.00 | 0.00 | 0.96 |
| W3 | 0.10 | 0.10 | 0.00 | 0.00 | 0.95 |
| W4 | 0.10 | 0.10 | 0.00 | 0.00 | 0.98 |
| Covariate1 | -0.40 | -0.40 | 0.00 | 0.00 | 0.95 |
| Covariate2 | 0.50 | 0.50 | 0.00 | 0.01 | 0.96 |
| Covariate3 | 0.20 | 0.20 | 0.00 | 0.00 | 0.96 |
| sigma | 1.00 | 1.00 | 0.00 | 0.63 | 0.96 |
| Simulation 3 |  |  |  |  |  |
| beta0_1 | 0.65 | 0.58 | -0.08 | 0.54 | 0.94 |
| beta0_2 | -0.38 | -0.28 | 0.10 | 0.34 | 0.93 |
| beta0_3 | 0.15 | 0.26 | 0.11 | 0.21 | 0.96 |
| beta0_4 | -0.98 | -0.80 | 0.18 | 0.57 | 0.91 |
| beta0_5 | 1.13 | 0.98 | -0.15 | 0.66 | 0.93 |
| beta1_1 | -1.70 | -1.66 | 0.04 | 0.11 | 0.93 |
| beta1_2 | -0.94 | -0.98 | -0.04 | 0.06 | 0.92 |
| beta1_3 | -0.11 | -0.16 | -0.05 | 0.03 | 0.95 |
| beta1_4 | -1.45 | -1.52 | -0.07 | 0.11 | 0.91 |
| beta1_5 | -1.88 | -1.80 | 0.08 | 0.14 | 0.92 |
| W1 | 0.30 | 0.29 | -0.01 | 0.00 | 0.90 |
| W2 | 0.45 | 0.44 | -0.01 | 0.00 | 0.93 |
| W3 | 0.24 | 0.23 | -0.01 | 0.00 | 0.92 |
| W4 | 0.01 | 0.04 | 0.03 | 0.01 | 0.94 |
| Covariate1 | -0.40 | -0.40 | 0.00 | 0.00 | 0.96 |
| Covariate2 | 0.50 | 0.50 | 0.00 | 0.01 | 0.96 |
| Covariate3 | 0.20 | 0.20 | 0.00 | 0.00 | 0.95 |
| sigma | 1.00 | 1.00 | 0.00 | 0.59 | 0.92 |
| Simulation 4 |  |  |  |  |  |
| beta0_1 | 0.65 | 0.63 | -0.02 | 0.15 | 0.95 |
| beta0_2 | -0.38 | -0.34 | 0.04 | 0.22 | 0.95 |
| beta0_3 | 0.15 | 0.23 | 0.08 | 0.19 | 0.96 |
| beta0_4 | -0.98 | -0.86 | 0.12 | 0.34 | 0.95 |
| beta0_5 | 1.13 | 0.90 | -0.23 | 0.32 | 0.93 |
| beta1_1 | 0.00 | 0.01 | 0.01 | 0.01 | 0.94 |
| beta1_2 | -0.94 | -0.96 | -0.02 | 0.03 | 0.95 |
| beta1_3 | -0.11 | -0.15 | -0.04 | 0.03 | 0.94 |
| beta1_4 | -1.45 | -1.50 | -0.05 | 0.06 | 0.94 |
| beta1_5 | -1.88 | -1.77 | 0.11 | 0.06 | 0.91 |
| W1 | 0.10 | 0.11 | 0.01 | 0.00 | 0.96 |
| W2 | 0.40 | 0.40 | 0.00 | 0.00 | 0.96 |
| W3 | 0.30 | 0.30 | -0.01 | 0.00 | 0.95 |
| W4 | 0.20 | 0.20 | -0.01 | 0.00 | 0.95 |
| Covariate1 | -0.40 | -0.40 | 0.00 | 0.00 | 0.94 |
| Covariate2 | 0.50 | 0.50 | 0.00 | 0.01 | 0.94 |
| Covariate3 | 0.20 | 0.20 | 0.00 | 0.00 | 0.95 |
| sigma | 1.00 | 1.00 | 0.00 | 0.58 | 0.95 |
| Simulation 5 |  |  |  |  |  |
| beta0_1 | 0.65 | 0.16 | -0.49 | 1.65 | 0.93 |
| beta0_2 | -0.38 | -0.01 | 0.38 | 1.59 | 0.94 |
| beta0_3 | 0.15 | 0.62 | 0.47 | 1.91 | 0.94 |
| beta0_4 | -0.98 | -0.41 | 0.57 | 1.88 | 0.93 |
| beta0_5 | 1.13 | 0.30 | -0.83 | 2.22 | 0.90 |
| beta1_1 | -1.70 | -1.48 | 0.22 | 0.15 | 0.92 |
| beta1_2 | -0.94 | -1.12 | -0.18 | 0.13 | 0.94 |
| beta1_3 | -0.11 | -0.40 | -0.29 | 0.23 | 0.92 |
| beta1_4 | -1.45 | -1.67 | -0.22 | 0.18 | 0.94 |
| beta1_5 | -1.88 | -1.48 | 0.40 | 0.27 | 0.81 |
| W1 | 0.10 | 0.13 | 0.03 | 0.01 | 0.98 |
| W2 | 0.40 | 0.39 | -0.01 | 0.01 | 0.97 |
| W3 | 0.30 | 0.29 | -0.01 | 0.01 | 0.97 |
| W4 | 0.20 | 0.20 | 0.00 | 0.01 | 0.97 |
| Covariate1 | -0.40 | -0.40 | 0.00 | 0.00 | 0.95 |
| Covariate2 | 0.50 | 0.50 | -0.01 | 0.05 | 0.93 |
| Covariate3 | 0.20 | 0.20 | 0.00 | 0.00 | 0.93 |
| sigma | 3.12 | 3.13 | 0.01 | 8.25 | 0.95 |
| Simulation 6 |  |  |  |  |  |
| beta0_1 | 0.65 | 0.49 | -0.16 | 0.18 | 0.93 |
| beta0_2 | -0.38 | -0.27 | 0.12 | 0.15 | 0.95 |
| beta0_3 | 0.15 | 0.22 | 0.07 | 0.14 | 0.95 |
| beta0_4 | -0.98 | -0.74 | 0.24 | 0.27 | 0.93 |
| beta0_5 | 1.13 | 0.81 | -0.32 | 0.29 | 0.89 |
| beta0_6 | -0.55 | -0.41 | 0.14 | 0.17 | 0.94 |
| beta0_7 | 0.98 | 0.87 | -0.11 | 0.19 | 0.96 |
| beta0_8 | 0.55 | 0.44 | -0.11 | 0.18 | 0.95 |
| beta0_9 | -0.18 | -0.18 | 0.00 | 0.15 | 0.96 |
| beta0_10 | 0.13 | 0.18 | 0.05 | 0.13 | 0.96 |
| beta1_1 | -1.70 | -1.62 | 0.08 | 0.03 | 0.91 |
| beta1_2 | -0.94 | -0.99 | -0.05 | 0.02 | 0.95 |
| beta1_3 | -0.11 | -0.14 | -0.03 | 0.02 | 0.97 |
| beta1_4 | -1.45 | -1.56 | -0.11 | 0.05 | 0.91 |
| beta1_5 | -1.88 | -1.72 | 0.16 | 0.05 | 0.86 |
| beta1_6 | -0.87 | -0.94 | -0.07 | 0.02 | 0.94 |
| beta1_7 | -0.04 | 0.01 | 0.05 | 0.03 | 0.96 |
| beta1_8 | -0.81 | -0.76 | 0.05 | 0.03 | 0.96 |
| beta1_9 | -1.95 | -1.95 | 0.00 | 0.02 | 0.95 |
| beta1_10 | -0.58 | -0.60 | -0.02 | 0.02 | 0.98 |
| W1 | 0.10 | 0.10 | 0.00 | 0.00 | 0.95 |
| W2 | 0.40 | 0.40 | 0.00 | 0.00 | 0.95 |
| W3 | 0.30 | 0.30 | 0.00 | 0.00 | 0.95 |
| W4 | 0.20 | 0.20 | 0.00 | 0.00 | 0.94 |
| Covariate1 | -0.40 | -0.40 | 0.00 | 0.00 | 0.94 |
| Covariate2 | 0.50 | 0.50 | 0.00 | 0.00 | 0.95 |
| Covariate3 | 0.20 | 0.20 | 0.00 | 0.00 | 0.94 |
| sigma | 0.80 | 0.80 | 0.00 | 0.32 | 0.93 |

**Table S2.** Mean estimate and standard deviation (SD) of the main parameters (intercept: beta_0_, slope: beta_1_, weights: W, model variance: sigma) of the six simulated scenarios using the Bayesian weighted quantile sum regression (BWQS).

| Parameters | Scenario 1 |  | Scenario 2 |  | Scenario 3 |  | Scenario 4 |  | Scenario 5 |  | Scenario 6 |  |
| --- | --- | --- | --- | --- | --- | --- | --- | --- | --- | --- | --- | --- |
|  | Mean Estimate | SD | Mean Estimate | SD | Mean Estimate | SD | Mean Estimate | SD | Mean Estimate | SD | Mean Estimate | SD |
| beta 0 | 0.15 | 0.78 | -0.16 | 1.52 | 0.31 | 0.76 | 0.23 | 0.77 | 0.15 | 1.17 | 0.04 | 0.67 |
| beta 1 | -1.29 | 0.31 | -1.14 | 0.73 | -1.36 | 0.31 | -0.78 | 0.20 | -1.30 | 0.28 | -1.05 | 0.21 |
| w1 | 0.12 | 0.10 | 0.62 | 0.22 | 0.28 | 0.06 | 0.14 | 0.08 | 0.14 | 0.07 | 0.12 | 0.08 |
| w2 | 0.39 | 0.07 | 0.13 | 0.14 | 0.43 | 0.08 | 0.39 | 0.09 | 0.38 | 0.09 | 0.39 | 0.07 |
| w3 | 0.29 | 0.06 | 0.13 | 0.12 | 0.23 | 0.06 | 0.28 | 0.08 | 0.28 | 0.08 | 0.29 | 0.06 |
| w4 | 0.20 | 0.06 | 0.12 | 0.12 | 0.06 | 0.08 | 0.19 | 0.07 | 0.20 | 0.08 | 0.19 | 0.06 |
| Covariate1 | -0.40 | 0.01 | -0.40 | 0.01 | -0.40 | 0.01 | -0.40 | 0.01 | -0.40 | 0.02 | -0.40 | 0.01 |
| Covariate2 | 0.50 | 0.10 | 0.51 | 0.11 | 0.51 | 0.10 | 0.50 | 0.15 | 0.49 | 0.23 | 0.50 | 0.11 |
| Covariate3 | 0.20 | 0.03 | 0.20 | 0.03 | 0.20 | 0.02 | 0.20 | 0.03 | 0.20 | 0.06 | 0.20 | 0.03 |
| sigma | 1.47 | 0.04 | 1.52 | 0.07 | 1.48 | 0.05 | 2.02 | 0.04 | 3.30 | 0.08 | 1.72 | 0.04 |

**Table S3.** Mean computing time in seconds (and Standard Deviation (SD)) for the iterations of the generated posterior parameter values of the Bayesian weighted quantile (BWQS) and the hierarchical BWQS (HBWQS) regressions across all simulated Scenarios.

| Computing Time in<br>seconds | BWQS |  | HBWQS |  |
| --- | --- | --- | --- | --- |
|  | Mean | SD | Mean | SD |
| Scenario 1 | 308 | 77 | 435 | 371 |
| Scenario 2 | 313 | 80 | 356 | 72 |
| Scenario 3 | 314 | 80 | 420 | 256 |
| Scenario 4 | 302 | 70 | 442 | 247 |
| Scenario 5 | 299 | 68 | 2413 | 2287 |
| Scenario 6 | 313 | 75 | 477 | 258 |

**Table S4.** Mean estimate, 95% and 80% Credible Intervals (CrI), number of effective sample size (ESS) and Rhat of the association (b1 coefficient) between the prenatal urinary metal mixture exposure (Arsenic (As), Cadmium (Cd), and Lead (Pb)) and gestational age in the PRISM New York City (NYC), PRISM Boston (BOS) and F1000 studies by using the Bayesian weighted quantile (BWQS) regression at individual sites and in the overall PRISM study aggregating all sites and by using the hierarchical BWQS (HBWQS) regression using all sites. Mean estimates, 95%CrI and 80%CrI, ESS, and Rhat of the intercept (b0 coefficient) and the contribution (weight (W)) of each metal to the mixture.

| Variable | Main analysis (n=1,349) |  |  |  |  | Including only women that delivered at term (n=1,276) |  |  |  |  |
| --- | --- | --- | --- | --- | --- | --- | --- | --- | --- | --- |
|  | Mean estimate | 95%CrI | 80%CrI | ESS | Rhat | Mean estimate | 95%CrI | 80%CrI | ESS | Rhat |
| HBWQS regression - Chain: 20000 - Burnin:10000 - Thinning: 10 |  |  |  |  |  |  |  |  |  |  |
| W - Arsenic | 0.21 | (0.01; 0.64) | (0.03; 0.45) | 804.98 | 1.00 | 0.34 | (0.02; 0.84) | (0.06; 0.66) | 949.02 | 1.00 |
| W - Cadmium | 0.64 | (0.12; 0.95) | (0.31; 0.89) | 875.90 | 1.00 | 0.32 | (0.01; 0.81) | (0.05; 0.65) | 999.99 | 1.00 |
| W - Lead | 0.15 | (0; 0.59) | (0.02; 0.35) | 975.74 | 1.00 | 0.34 | (0.01; 0.83) | (0.05; 0.68) | 1029.47 | 1.00 |
| b0 - NYC | 40.42 | (39; 41.79) | (39.56; 41.3) | 988.37 | 1.00 | 40.29 | (39.53; 41.1) | (39.78; 40.81) | 1002.66 | 1.00 |
| b0 - BOS | 40.46 | (38.98; 41.9) | (39.58; 41.35) | 1007.15 | 1.00 | 40.31 | (39.54; 41.12) | (39.79; 40.84) | 1015.25 | 1.00 |
| b0 - F1000 | 40.45 | (39; 41.81) | (39.57; 41.32) | 991.65 | 1.00 | 40.29 | (39.53; 41.11) | (39.78; 40.82) | 994.24 | 1.00 |
| b1 - NYC | -0.22 | **(-0.4; -0.02)** | **(-0.34; -0.1)** | 768.49 | 1.01 | -0.02 | (-0.13; 0.08) | (-0.09; 0.04) | 958.99 | 1.00 |
| b1 - BOS | -0.01 | (-0.21; 0.16) | (-0.13; 0.1) | 838.59 | 1.00 | 0.03 | (-0.08; 0.14) | (-0.04; 0.1) | 905.78 | 1.00 |
| b1 - F1000 | 0.01 | (-0.16; 0.16) | (-0.09; 0.1) | 941.53 | 1.00 | 0.01 | (-0.08; 0.11) | (-0.05; 0.08) | 970.94 | 1.00 |
| BWQS in the overall population, combining all studies - Chain: 10000 - Burnin:5000 - Thinning: 3 |  |  |  |  |  |  |  |  |  |  |
| W - Arsenic | 0.26 | (0; 0.72) | (0.03; 0.56) | 1585.21 | 1.00 | 0.32 | (0.01; 0.83) | (0.05; 0.65) | 1484.94 | 1.00 |
| W - Cadmium | 0.47 | (0.03; 0.9) | (0.12; 0.81) | 1643.43 | 1.00 | 0.35 | (0.01; 0.85) | (0.06; 0.7) | 1488.90 | 1.00 |
| W - Lead | 0.27 | (0.01; 0.76) | (0.04; 0.56) | 1605.99 | 1.00 | 0.33 | (0.01; 0.84) | (0.04; 0.68) | 1590.84 | 1.00 |
| b0 | 40.50 | (39.14; 41.82) | (39.62; 41.38) | 1498.34 | 1.00 | 40.26 | (39.49; 41.04) | (39.75; 40.76) | 1409.55 | 1.00 |
| b1 | -0.09 | (-0.24; 0.06) | (-0.19; 0.01) | 1462.35 | 1.00 | 0.00 | (-0.09; 0.09) | (-0.06; 0.05) | 1571.83 | 1.00 |
| BWQS in the PRISM NYC study- Chain: 10000 - Burnin:5000 - Thinning: 3 |  |  |  |  |  |  |  |  |  |  |
| W - Arsenic | 0.27 | (0.01; 0.75) | (0.04; 0.58) | 1559.34 | 1.00 | 0.34 | (0.02; 0.83) | (0.06; 0.67) | 1686.29 | 1.00 |
| W - Cadmium | 0.44 | (0.02; 0.91) | (0.07; 0.81) | 1431.38 | 1.00 | 0.33 | (0.01; 0.83) | (0.05; 0.66) | 1615.16 | 1.00 |
| W - Lead | 0.29 | (0.01; 0.83) | (0.04; 0.63) | 1545.03 | 1.00 | 0.33 | (0.01; 0.84) | (0.05; 0.67) | 1490.51 | 1.00 |
| b0 | 42.53 | (38.69; 46.29) | (39.92; 45.07) | 1169.48 | 1.00 | 41.21 | (39.31; 43.25) | (39.95; 42.5) | 1430.98 | 1.00 |
| b1 | -0.11 | (-0.53; 0.32) | (-0.4; 0.17) | 1264.61 | 1.00 | -0.02 | (-0.22; 0.19) | (-0.15; 0.12) | 1468.16 | 1.00 |
| BWQS in the PRISM BOS study - Chain: 10000 - Burnin:5000 - Thinning: 3 |  |  |  |  |  |  |  |  |  |  |
| W - Arsenic | 0.32 | (0.01; 0.82) | (0.05; 0.66) | 1591.48 | 1.00 | 0.29 | (0.01; 0.79) | (0.04; 0.61) | 1774.90 | 1.00 |
| W - Cadmium | 0.34 | (0.01; 0.83) | (0.05; 0.68) | 1601.25 | 1.00 | 0.34 | (0.02; 0.83) | (0.06; 0.67) | 1721.96 | 1.00 |
| W - Lead | 0.34 | (0.02; 0.82) | (0.06; 0.67) | 1486.44 | 1.00 | 0.37 | (0.01; 0.88) | (0.07; 0.72) | 1629.02 | 1.00 |
| b0 | 40.91 | (38.17; 43.63) | (39.14; 42.74) | 1566.84 | 1.00 | 39.86 | (37.73; 42.05) | (38.43; 41.31) | 1389.18 | 1.00 |
| b1 | 0.02 | (-0.23; 0.28) | (-0.15; 0.19) | 1592.24 | 1.00 | 0.08 | (-0.11; 0.27) | (-0.05; 0.21) | 1527.69 | 1.00 |
| BWQS in the F1000 study - Chain: 10000 - Burnin:5000 - Thinning: 3 |  |  |  |  |  |  |  |  |  |  |
| W - Arsenic | 0.32 | (0.01; 0.82) | (0.05; 0.65) | 1683.58 | 1.00 | 0.31 | (0.01; 0.82) | (0.05; 0.64) | 1789.44 | 1.00 |
| W - Cadmium | 0.36 | (0.01; 0.83) | (0.07; 0.69) | 1613.55 | 1.00 | 0.35 | (0.02; 0.84) | (0.07; 0.69) | 1721.42 | 1.00 |
| W - Lead | 0.31 | (0.01; 0.83) | (0.05; 0.65) | 1686.19 | 1.00 | 0.34 | (0.02; 0.84) | (0.06; 0.68) | 1928.83 | 1.00 |
| b0 | 39.80 | (38.64; 40.91) | (39.09; 40.54) | 1656.90 | 1.00 | 40.03 | (39.03; 41.03) | (39.38; 40.67) | 1452.24 | 1.00 |
| b1 | -0.05 | (-0.18; 0.08) | (-0.13; 0.03) | 1508.71 | 1.00 | -0.02 | (-0.13; 0.1) | (-0.09; 0.06) | 1547.28 | 1.00 |

**Table S5.**
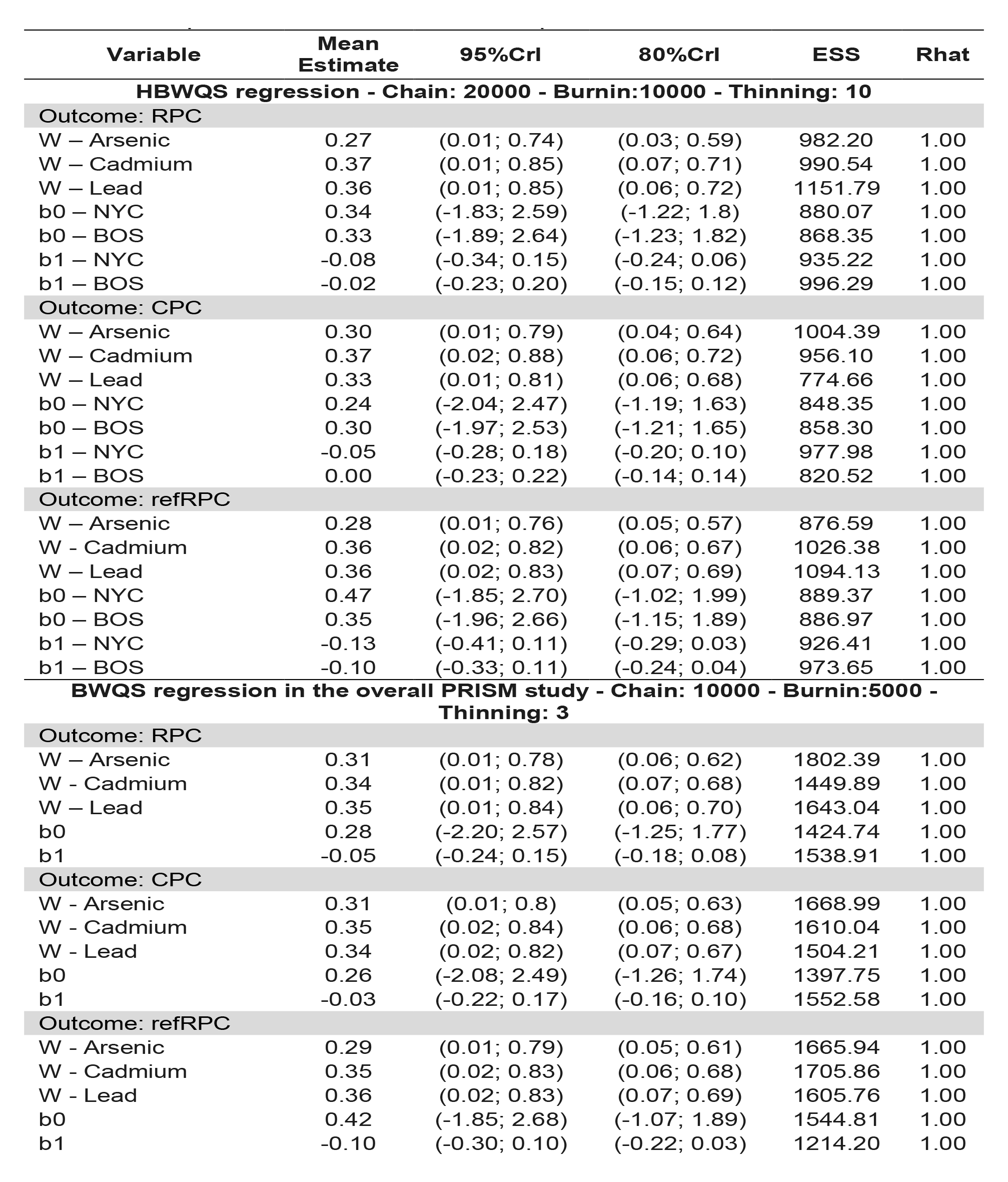
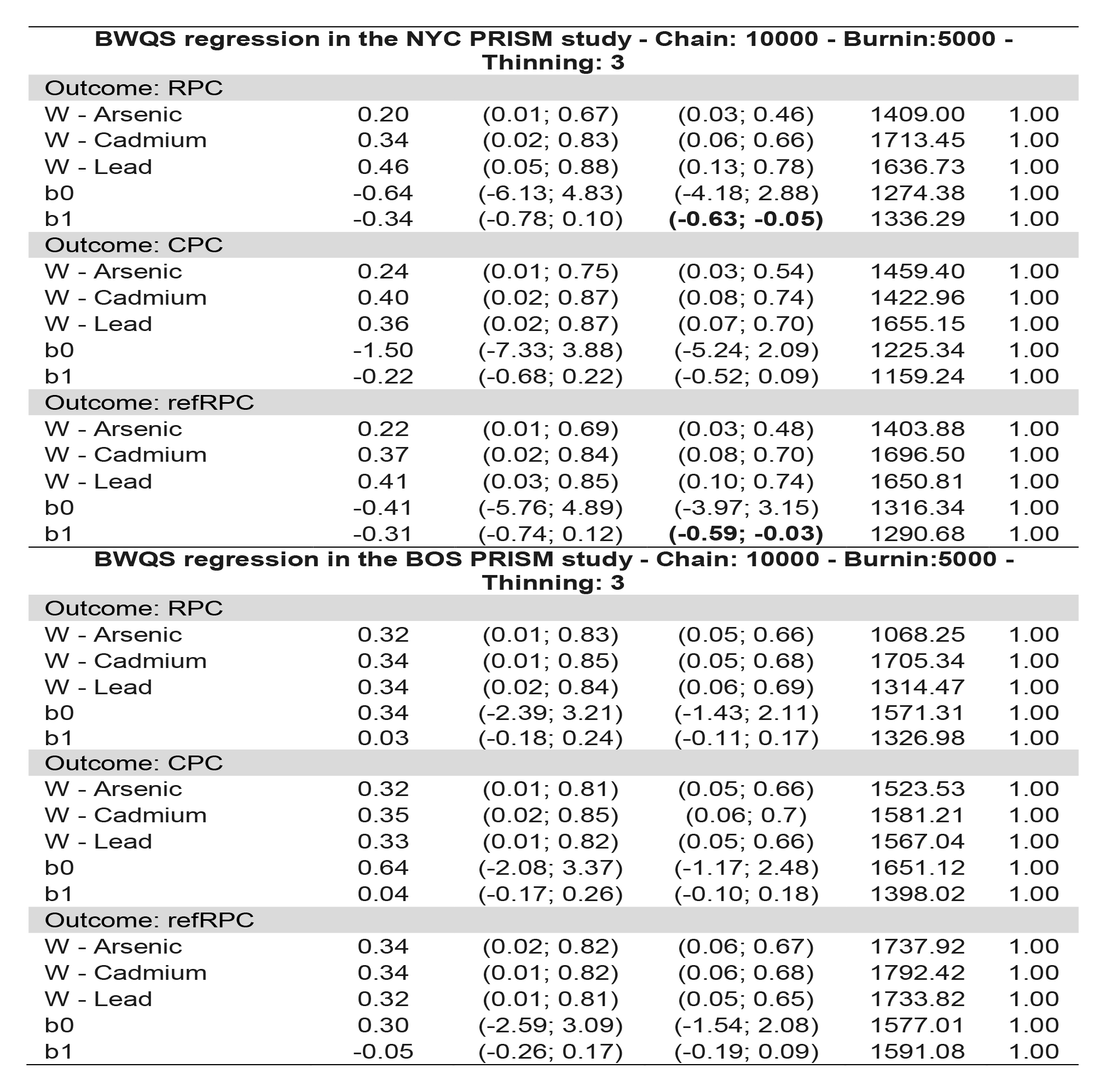
Mean estimates, 95% and 80% Credible Intervals (CrI), number of effective sample size (ESS) and Rhat of the association (b1 coefficient) between the prenatal urinary metal mixture exposure (Arsenic (As), Cadmium (Cd), and Lead (Pb)) and gestational age acceleration metrics (RPC, CPC, refRPC) (weeks) in the PRISM New York City (NYC), and PRISM Boston (BOS) by using the Bayesian weighted quantile (BWQS) regression at individual sites and in the overall PRISM study aggregating all sites and by using the hierarchical BWQS (HBWQS) regression using both sites. Mean estimates, 95%CrI and 80%CrI, ESS, and Rhat of the intercept (b0 coefficient) and the contribution (weight (W)) of each metal to the mixture. RPC= robust placental clock; CPC: control placental clock; refRPC: refined RPC

| Variable | Mean Estimate | 95%CrI | 80%CrI | ESS | Rhat |
| --- | --- | --- | --- | --- | --- |
| **HBWQS regression - Chain: 20000 - Burnin:10000 - Thinning: 10** |  |  |  |  |  |
| **Outcome: RPC** |  |  |  |  |  |
| W – Arsenic | 0.27 | (0.01; 0.74) | (0.03; 0.59) | 982.20 | 1.00 |
| W – Cadmium | 0.37 | (0.01; 0.85) | (0.07; 0.71) | 990.54 | 1.00 |
| W – Lead | 0.36 | (0.01; 0.85) | (0.06; 0.72) | 1151.79 | 1.00 |
| b0 – NYC | 0.34 | (-1.83; 2.59) | (-1.22; 1.8) | 880.07 | 1.00 |
| b0 – BOS | 0.33 | (-1.89; 2.64) | (-1.23; 1.82) | 868.35 | 1.00 |
| b1 – NYC | -0.08 | (-0.34; 0.15) | (-0.24; 0.06) | 935.22 | 1.00 |
| b1 – BOS | -0.02 | (-0.23; 0.20) | (-0.15; 0.12) | 996.29 | 1.00 |
| **Outcome: CPC** |  |  |  |  |  |
| W – Arsenic | 0.30 | (0.01; 0.79) | (0.04; 0.64) | 1004.39 | 1.00 |
| W – Cadmium | 0.37 | (0.02; 0.88) | (0.06; 0.72) | 956.10 | 1.00 |
| W – Lead | 0.33 | (0.01; 0.81) | (0.06; 0.68) | 774.66 | 1.00 |
| b0 – NYC | 0.24 | (-2.04; 2.47) | (-1.19; 1.63) | 848.35 | 1.00 |
| b0 – BOS | 0.30 | (-1.97; 2.53) | (-1.21; 1.65) | 858.30 | 1.00 |
| b1 – NYC | -0.05 | (-0.28; 0.18) | (-0.20; 0.10) | 977.98 | 1.00 |
| b1 – BOS | 0.00 | (-0.23; 0.22) | (-0.14; 0.14) | 820.52 | 1.00 |
| **Outcome: refRPC** |  |  |  |  |  |
| W – Arsenic | 0.28 | (0.01; 0.76) | (0.05; 0.57) | 876.59 | 1.00 |
| W – Cadmium | 0.36 | (0.02; 0.82) | (0.06; 0.67) | 1026.38 | 1.00 |
| W – Lead | 0.36 | (0.02; 0.83) | (0.07; 0.69) | 1094.13 | 1.00 |
| b0 – NYC | 0.47 | (-1.85; 2.70) | (-1.02; 1.99) | 889.37 | 1.00 |
| b0 – BOS | 0.35 | (-1.96; 2.66) | (-1.15; 1.89) | 886.97 | 1.00 |
| b1 – NYC | -0.13 | (-0.41; 0.11) | (-0.29; 0.03) | 926.41 | 1.00 |
| b1 – BOS | -0.10 | (-0.33; 0.11) | (-0.24; 0.04) | 973.65 | 1.00 |
| **BWQS regression in the overall PRISM study - Chain: 10000 - Burnin:5000 - Thinning: 3** |  |  |  |  |  |
| **Outcome: RPC** |  |  |  |  |  |
| W – Arsenic | 0.31 | (0.01; 0.78) | (0.06; 0.62) | 1802.39 | 1.00 |
| W – Cadmium | 0.34 | (0.01; 0.82) | (0.07; 0.68) | 1449.89 | 1.00 |
| W – Lead | 0.35 | (0.01; 0.84) | (0.06; 0.70) | 1643.04 | 1.00 |
| b0 | 0.28 | (-2.20; 2.57) | (-1.25; 1.77) | 1424.74 | 1.00 |
| b1 | -0.05 | (-0.24; 0.15) | (-0.18; 0.08) | 1538.91 | 1.00 |
| **Outcome: CPC** |  |  |  |  |  |
| W – Arsenic | 0.31 | (0.01; 0.8) | (0.05; 0.63) | 1668.99 | 1.00 |
| W – Cadmium | 0.35 | (0.02; 0.84) | (0.06; 0.68) | 1610.04 | 1.00 |
| W – Lead | 0.34 | (0.02; 0.82) | (0.07; 0.67) | 1504.21 | 1.00 |
| b0 | 0.26 | (-2.08; 2.49) | (-1.26; 1.74) | 1397.75 | 1.00 |
| b1 | -0.03 | (-0.22; 0.17) | (-0.16; 0.10) | 1552.58 | 1.00 |
| **Outcome: refRPC** |  |  |  |  |  |
| W – Arsenic | 0.29 | (0.01; 0.79) | (0.05; 0.61) | 1665.94 | 1.00 |
| W – Cadmium | 0.35 | (0.02; 0.83) | (0.06; 0.68) | 1705.86 | 1.00 |
| W – Lead | 0.36 | (0.02; 0.83) | (0.07; 0.69) | 1605.76 | 1.00 |
| b0 | 0.42 | (-1.85; 2.68) | (-1.07; 1.89) | 1544.81 | 1.00 |
| b1 | -0.10 | (-0.30; 0.10) | (-0.22; 0.03) | 1214.20 | 1.00 |

| **BWQS regression in the NYC PRISM study - Chain: 10000 - Burnin:5000 - Thinning: 3** |  |  |  |  |  |
| --- | --- | --- | --- | --- | --- |
| **Outcome: RPC** |  |  |  |  |  |
| W - Arsenic | 0.20 | (0.01; 0.67) | (0.03; 0.46) | 1409.00 | 1.00 |
| W - Cadmium | 0.34 | (0.02; 0.83) | (0.06; 0.66) | 1713.45 | 1.00 |
| W - Lead | 0.46 | (0.05; 0.88) | (0.13; 0.78) | 1636.73 | 1.00 |
| b0 | -0.64 | (-6.13; 4.83) | (-4.18; 2.88) | 1274.38 | 1.00 |
| b1 | -0.34 | (-0.78; 0.10) | **(-0.63; -0.05)** | 1336.29 | 1.00 |
| **Outcome: CPC** |  |  |  |  |  |
| W - Arsenic | 0.24 | (0.01; 0.75) | (0.03; 0.54) | 1459.40 | 1.00 |
| W - Cadmium | 0.40 | (0.02; 0.87) | (0.08; 0.74) | 1422.96 | 1.00 |
| W - Lead | 0.36 | (0.02; 0.87) | (0.07; 0.70) | 1655.15 | 1.00 |
| b0 | -1.50 | (-7.33; 3.88) | (-5.24; 2.09) | 1225.34 | 1.00 |
| b1 | -0.22 | (-0.68; 0.22) | (-0.52; 0.09) | 1159.24 | 1.00 |
| **Outcome: refRPC** |  |  |  |  |  |
| W - Arsenic | 0.22 | (0.01; 0.69) | (0.03; 0.48) | 1403.88 | 1.00 |
| W - Cadmium | 0.37 | (0.02; 0.84) | (0.08; 0.70) | 1696.50 | 1.00 |
| W - Lead | 0.41 | (0.03; 0.85) | (0.10; 0.74) | 1650.81 | 1.00 |
| b0 | -0.41 | (-5.76; 4.89) | (-3.97; 3.15) | 1316.34 | 1.00 |
| b1 | -0.31 | (-0.74; 0.12) | **(-0.59; -0.03)** | 1290.68 | 1.00 |
| **BWQS regression in the BOS PRISM study - Chain: 10000 - Burnin:5000 - Thinning: 3** |  |  |  |  |  |
| **Outcome: RPC** |  |  |  |  |  |
| W - Arsenic | 0.32 | (0.01; 0.83) | (0.05; 0.66) | 1068.25 | 1.00 |
| W - Cadmium | 0.34 | (0.01; 0.85) | (0.05; 0.68) | 1705.34 | 1.00 |
| W - Lead | 0.34 | (0.02; 0.84) | (0.06; 0.69) | 1314.47 | 1.00 |
| b0 | 0.34 | (-2.39; 3.21) | (-1.43; 2.11) | 1571.31 | 1.00 |
| b1 | 0.03 | (-0.18; 0.24) | (-0.11; 0.17) | 1326.98 | 1.00 |
| **Outcome: CPC** |  |  |  |  |  |
| W - Arsenic | 0.32 | (0.01; 0.81) | (0.05; 0.66) | 1523.53 | 1.00 |
| W - Cadmium | 0.35 | (0.02; 0.85) | (0.06; 0.7) | 1581.21 | 1.00 |
| W - Lead | 0.33 | (0.01; 0.82) | (0.05; 0.66) | 1567.04 | 1.00 |
| b0 | 0.64 | (-2.08; 3.37) | (-1.17; 2.48) | 1651.12 | 1.00 |
| b1 | 0.04 | (-0.17; 0.26) | (-0.10; 0.18) | 1398.02 | 1.00 |
| **Outcome: refRPC** |  |  |  |  |  |
| W - Arsenic | 0.34 | (0.02; 0.82) | (0.06; 0.67) | 1737.92 | 1.00 |
| W - Cadmium | 0.34 | (0.01; 0.82) | (0.06; 0.68) | 1792.42 | 1.00 |
| W - Lead | 0.32 | (0.01; 0.81) | (0.05; 0.65) | 1733.82 | 1.00 |
| b0 | 0.30 | (-2.59; 3.09) | (-1.54; 2.08) | 1577.01 | 1.00 |
| b1 | -0.05 | (-0.26; 0.17) | (-0.19; 0.09) | 1591.08 | 1.00 |

**Table S6.** Computing time (in seconds) for 1,000 generated posterior parameter values from the Bayesian weighted quantile (BWQS) and the hierarchical BWQS (HBWQS) regressions during the warm-up and the post warm-up periods in the real case scenarios.

| Outcome | BWQS |  | HBWQS |  |
| --- | --- | --- | --- | --- |
|  | Warm-up | Post Warm-up | Warm-up | Post Warm-up |
| Gestational age | 71 | 47 | 2811 | 783 |
| RPC | 32 | 38 | 676 | 574 |
| CPC | 36 | 45 | 603 | 491 |
| refRPC | 35 | 55 | 657 | 469 |

RPC= robust placental clock; CPC: control placental clock; refRPC: refined RPC

## References

1. Niedzwiecki MM, Miller GW. The Exposome Paradigm in Human Health: Lessons from the Emory Exposome Summer Course. Environ Health Perspect 2017;125(6):064502. (In eng). DOI: 10.1289/ehp1712.

2. Taylor KW, Joubert BR, Braun JM, et al. Statistical Approaches for Assessing Health Effects of Environmental Chemical Mixtures in Epidemiology: Lessons from an Innovative Workshop. Environmental Health Perspectives 2016;124(12):A227–A229. DOI: doi:10.1289/EHP547.

3. Gibson EA, Goldsmith J, Kioumourtzoglou M-A. Complex Mixtures, Complex Analyses: an Emphasis on Interpretable Results. Curr Environ Health Rep 2019 (In eng). DOI: 10.1007/s40572-019-00229-5.

4. Becker J-M, Ringle CM, Sarstedt M, Völckner F. How collinearity affects mixture regression results. Marketing Letters 2015;26(4):643–659. (journal article). DOI: 10.1007/s11002-014-9299-9.

5. Yorita Christensen KL, White P. A methodological approach to assessing the health impact of environmental chemical mixtures: PCBs and hypertension in the National Health and Nutrition Examination Survey. International journal of environmental research and public health 2011;8(11):4220–37. (In eng). DOI: 10.3390/ijerph8114220.

6. Valeri L, Mazumdar MM, Bobb JF, et al. The Joint Effect of Prenatal Exposure to Metal Mixtures on Neurodevelopmental Outcomes at 20-40 Months of Age: Evidence from Rural Bangladesh. Environ Health Perspect 2017;125(6):067015. (In eng). DOI: 10.1289/ehp614.

7. Stafoggia M, Breitner S, Hampel R, Basagana X. Statistical Approaches to Address Multi-Pollutant Mixtures and Multiple Exposures: the State of the Science. Curr Environ Health Rep 2017;4(4):481–490. (In eng). DOI: 10.1007/s40572-017-0162-z.

8. Kirk P, Griffin JE, Savage RS, Ghahramani Z, Wild DL. Bayesian correlated clustering to integrate multiple datasets. Bioinformatics (Oxford, England) 2012;28(24):3290–7. (In eng). DOI: 10.1093/bioinformatics/bts595.

9. Gottfredson NC, Cole VT, Giordano ML, Bauer DJ, Hussong AM, Ennett ST. Simplifying the implementation of modern scale scoring methods with an automated R package: Automated moderated nonlinear factor analysis (aMNLFA). Addictive Behaviors 2019;94:65–73. DOI: https://doi.org/10.1016/j.addbeh.2018.10.031.

10. De Vito R, Bellio R, Trippa L, Parmigiani G. Bayesian multistudy factor analysis for high-throughput biological data. The Annals of Applied Statistics 2021;15(4):1723–1741, 19. (https://doi.org/10.1214/21-AOAS1456).

11. Colicino E, Foppa Pedretti N, Busgang S, Gennings C. Per-and poly-fluoroalkyl substances and bone mineral density: results from the Bayesian weighted quantile sum regression. medRxiv 2019:19010710. DOI: 10.1101/19010710.

12. Gennings C, Carrico C, Factor-Litvak P, Krigbaum N, Cirillo PM, Cohn BA. A cohort study evaluation of maternal PCB exposure related to time to pregnancy in daughters. Environ Health 2013;12(1):66. DOI: 10.1186/1476-069X-12-66.

13. Czarnota J, Gennings C, Wheeler DC. Assessment of weighted quantile sum regression for modeling chemical mixtures and cancer risk. Cancer informatics 2015;14(Suppl 2):159–71. (In eng). DOI: 10.4137/cin.s17295.

14. Carrico C, Gennings C, Wheeler DC, Factor-Litvak P. Characterization of Weighted Quantile Sum Regression for Highly Correlated Data in a Risk Analysis Setting. Journal of Agricultural, Biological, and Environmental Statistics 2015;20(1):100–120. (journal article). DOI: 10.1007/s13253-014-0180-3.

15. Colicino E, Pedretti NF, Busgang SA, Gennings C. Per-and poly-fluoroalkyl substances and bone mineral density: Results from the Bayesian weighted quantile sum regression. Environ Epidemiol 2020;4(3):e092. (In eng). DOI: 10.1097/ee9.0000000000000092.

16. Watanabe K, Taskesen E, van Bochoven A, Posthuma D. Functional mapping and annotation of genetic associations with FUMA. Nature Communications 2017;8:1826. DOI: 10.1038/s41467-017-01261-5.

17. SW. Asymptotic Equivalence of Bayes Cross Validation and Widely Applicable Information Criterion in Singular Learning Theory. The Journal of Machine Learning Research archive 2010;11.

18. Zhang X, Liu SH, Geron M, et al. Prenatal exposure to PM(2.5) and childhood cognition: Accounting for between-site heterogeneity in a pooled analysis of ECHO cohorts in the Northeastern United States. Environ Res 2022;214(Pt 4):114163. (In eng). DOI: 10.1016/j.envres.2022.114163.

19. Cowell W, Colicino E, Tanner E, et al. Prenatal toxic metal mixture exposure and newborn telomere length: Modification by maternal antioxidant intake. Environmental Research 2020;190:110009. DOI: https://doi.org/10.1016/j.envres.2020.110009.

20. Hazrati S, Wong WSW, Huddleston K, et al. Clinical, Social, and Genetic Factors Associated with Obesity at 12 Months of Age. The Journal of pediatrics 2018;196:175–181.e7. (In eng). DOI: 10.1016/j.jpeds.2017.12.042.

21. Hazrati S, Hourigan SK, Waller A, et al. Investigating the accuracy of parentally reported weights and lengths at 12 months of age as compared to measured weights and lengths in a longitudinal childhood genome study. BMJ open 2016;6(8):e011653. (In eng). DOI: 10.1136/bmjopen-2016-011653.

22. Blaisdell CJ, Park C, Hanspal M, et al. The NIH ECHO Program: investigating how early environmental influences affect child health. Pediatr Res 2022;92(5):1215–1216. (In eng). DOI: 10.1038/s41390-021-01574-8.

23. Gillman MW, Blaisdell CJ. Environmental influences on Child Health Outcomes, a Research Program of the National Institutes of Health. Curr Opin Pediatr 2018;30(2):260–262. (In eng). DOI: 10.1097/mop.0000000000000600.

24. Camerota M, Bollen KA. Birth Weight, Birth Length, and Gestational Age as Indicators of Favorable Fetal Growth Conditions in a US Sample. PLoS One 2016;11(4):e0153800. (In eng). DOI: 10.1371/journal.pone.0153800.

25. Basso O, Wilcox AJ, Weinberg CR. Birth weight and mortality: causality or confounding? American journal of epidemiology 2006;164(4):303–11. (In eng). DOI: 10.1093/aje/kwj237.

26. Rosa MJ, Pajak A, Just AC, et al. Prenatal exposure to PM2.5 and birth weight: A pooled analysis from three North American longitudinal pregnancy cohort studies. Environment international 2017;107:173–180. (In eng). DOI: 10.1016/j.envint.2017.07.012.

27. Hoffman CS, Messer LC, Mendola P, Savitz DA, Herring AH, Hartmann KE. Comparison of gestational age at birth based on last menstrual period and ultrasound during the first trimester. Paediatric and perinatal epidemiology 2008;22(6):587–96. (In eng). DOI: 10.1111/j.1365-3016.2008.00965.x.

28. Saigal S, Doyle LW. An overview of mortality and sequelae of preterm birth from infancy to adulthood. Lancet (London, England) 2008;371(9608):261-9. (In eng). DOI: 10.1016/s0140-6736(08)60136-1.

29. Russell RB, Green NS, Steiner CA, et al. Cost of hospitalization for preterm and low birth weight infants in the United States. Pediatrics 2007;120(1):e1–9. (In eng). DOI: 10.1542/peds.2006-2386.

30. Lee Y, Choufani S, Weksberg R, et al. Placental epigenetic clocks: estimating gestational age using placental DNA methylation levels. Aging (Albany NY) 2019;11(12):4238–4253. (In eng). DOI: 10.18632/aging.102049.

31. Bozack AK, Colicino E, Just AC, et al. Associations between infant sex and DNA methylation across umbilical cord blood, artery, and placenta samples. Epigenetics 2022;17(10):1080–1097. (In eng). DOI: 10.1080/15592294.2021.1985300.

32. Janssen BG, Byun H-M, Gyselaers W, Lefebvre W, Baccarelli AA, Nawrot TS. Placental mitochondrial methylation and exposure to airborne particulate matter in the early life environment: an ENVIR ON AGE birth cohort study. Epigenetics 2015;10(6):536–544.

33. De Carli MM, Baccarelli AA, Trevisi L, et al. Epigenome-wide cross-tissue predictive modeling and comparison of cord blood and placental methylation in a birth cohort. Epigenomics 2017;9(3):231–240. (In eng). DOI: 10.2217/epi-2016-0109.

34. Brunst KJ, Tignor N, Just A, et al. Cumulative lifetime maternal stress and epigenome-wide placental DNA methylation in the PRISM cohort. Epigenetics 2018;13(6):665–681.

35. Heiss JA, Just AC. Identifying mislabeled and contaminated DNA methylation microarray data: an extended quality control toolset with examples from GEO. Clinical Epigenetics 2018;10(1):1–9.

36. Heiss JA, Just AC. Improved filtering of DNA methylation microarray data by detection p values and its impact on downstream analyses. Clinical Epigenetics 2019;11(1):1–8.

37. Xu Z, Langie SA, De Boever P, Taylor JA, Niu L. RELIC: a novel dye-bias correction method for Illumina Methylation BeadChip. BMC genomics 2017;18(1):1–7.

38. Geron M, Cowell W, Amarasiriwardena C, et al. Racial/ethnic and neighborhood disparities in metals exposure during pregnancy in the Northeastern United States. Science of The Total Environment 2022;820:153249. DOI: https://doi.org/10.1016/j.scitotenv.2022.153249.

39. Brooks S, Gelman A, Jones G, Meng X-L. Handbook of markov chain monte carlo: CRC press, 2011.

40. Gelman A, Carlin JB, Stern HS, Rubin DB. Bayesian data analysis: Chapman and Hall/CRC, 1995.

41. Raudenbush SW, Bryk AS. Hierarchical linear models: Applications and data analysis methods: sage, 2002.

42. Gelman A, Hill J. Data analysis using regression and multilevel/hierarchical models: Cambridge university press, 2006.

43. 43. McElreath R. Statistical rethinking: A Bayesian course with examples in R and Stan: Chapman and Hall/CRC, 2020.

44. Curran PJ, Hussong AM. Integrative data analysis: the simultaneous analysis of multiple data sets. Psychol Methods 2009;14(2):81–100. (In eng). DOI: 10.1037/a0015914.

45. Claus Henn B, Ettinger AS, Hopkins MR, et al. Prenatal Arsenic Exposure and Birth Outcomes among a Population Residing near a Mining-Related Superfund Site. Environ Health Perspect 2016;124(8):1308–15. DOI: 10.1289/ehp.1510070.

46. Kile ML, Cardenas A, Rodrigues E, et al. Estimating Effects of Arsenic Exposure During Pregnancy on Perinatal Outcomes in a Bangladeshi Cohort. Epidemiology (Cambridge, Mass) 2016;27(2):173–81. (In eng). DOI: 10.1097/ede.0000000000000416.

47. Rahman ML, Valeri L, Kile ML, et al. Investigating causal relation between prenatal arsenic exposure and birthweight: Are smaller infants more susceptible? Environment international 2017;108:32–40. DOI: 10.1016/j.envint.2017.07.026.

48. Rahman ML, Oken E, Hivert MF, et al. Early pregnancy exposure to metal mixture and birth outcomes - A prospective study in Project Viva. Environ Int 2021;156:106714. (In eng). DOI: 10.1016/j.envint.2021.106714.

49. Eum JH, Cheong HK, Ha EH, et al. Maternal blood manganese level and birth weight: a MOCEH birth cohort study. Environmental health: a global access science source 2014;13(1):31. (In eng). DOI: 10.1186/1476-069x-13-31.

50. Perkins M, Wright RO, Amarasiriwardena CJ, Jayawardene I, Rifas-Shiman SL, Oken E. Very low maternal lead level in pregnancy and birth outcomes in an eastern Massachusetts population. Annals of epidemiology 2014;24(12):915–9. (In eng). DOI: 10.1016/j.annepidem.2014.09.007.

51. Rahman ML, Kile ML, Rodrigues EG, et al. Prenatal arsenic exposure, child marriage, and pregnancy weight gain: Associations with preterm birth in Bangladesh. Environment international 2018;112:23–32. DOI: 10.1016/j.envint.2017.12.004.

52. Howe CG, Henn BC, Eckel SP, et al. Prenatal Metal Mixtures and Birth Weight for Gestational Age in a Predominately Lower-Income Hispanic Pregnancy Cohort in Los Angeles. Environmental Health Perspectives 2020;128(11):117001. DOI: doi:10.1289/EHP7201.

53. Howe CG, Nozadi SS, Garcia E, et al. Prenatal metal(loid) mixtures and birth weight for gestational age: A pooled analysis of three cohorts participating in the ECHO program. Environment International 2022;161:107102. DOI: https://doi.org/10.1016/j.envint.2022.107102.

54. Luo Y, McCullough LE, Tzeng J-Y, et al. Maternal blood cadmium, lead and arsenic levels, nutrient combinations, and offspring birthweight. BMC Public Health 2017;17(1):354. DOI: 10.1186/s12889-017-4225-8.

55. Lee M-S, Eum K-D, Golam M, et al. Umbilical Cord Blood Metal Mixtures and Birth Size in Bangladeshi Children. Environmental Health Perspectives 2021;129(5):057006. DOI: doi:10.1289/EHP7502.

56. Pohl HR, van Engelen JG, Wilson J, Sips AJ. Risk assessment of chemicals and pharmaceuticals in the pediatric population: a workshop report. Regulatory toxicology and pharmacology: RTP 2005;42(1):83–95. (In eng). DOI: 10.1016/j.yrtph.2005.01.005.

57. Braun JM, Gennings C, Hauser R, Webster TF. What Can Epidemiological Studies Tell Us about the Impact of Chemical Mixtures on Human Health? Environmental Health Perspectives 2016;124(1):A6–A9. DOI: doi:10.1289/ehp.1510569.

58. Holland P, Dorans N, Brennan R. Educational measurement. Praeger Publishers, Westport, CT; 2006.

59. Kreft I, de Leeuw J. Introducing Multilevel Modeling. London 1998.

60. Kiecolt KJ, Brinberg D, Auspurg K, Nathan LE, Nathan LE. Secondary analysis of survey data: Sage, 1985.

